# Mutations in 329 probands with suspected renal electrolyte disorders

**DOI:** 10.1101/2025.04.28.25326317

**Authors:** Renan Eduardo Aparicio, Sheng Chih Jin, Weilai Dong, Samir Zaidi, Michael C Sierant, James Knight, Robert D Bjornson, Christopher Castaldi, Shrikant M Mane, Thomas M. Kaneko, Carol Nelson-Williams, Richard P. Lifton

## Abstract

The spectrum of coding and non-coding of mutations that contribute to Mendelian diseases is largely unknown. This question is broadly relevant to molecular diagnostic efforts in real-world settings, particularly when the scope of biochemical and other tests performed may be limited prior to genetic testing. We report the results of whole exome sequencing of DNA from 329 patients referred by physicians for suspicion of genetic causes of renal electrolyte disorders, predominantly featuring hypokalemia and/or hypomagnesemia with normal to low blood pressure. 295 probands were found to have known/likely disease-causing genotypes; these included 254 with biallelic mutations in one of 9 genes known to cause salt wasting, and 34 with monoallelic mutations in these same genes. Seven other probands had mutations in genes known to cause renal hypomagnesemia. Among these, 197 probands had biallelic or monoallelic (n = 7) damaging variants in the Gitelman syndrome (GS) gene *SLC12A3*, which encodes the Na-Cl cotransporter of the distal convoluted tubule. WGS sequencing in the 7 monoallelic probands, who all had pathognomonic electrolyte values for GS, identified two deep intronic variants that introduced splice sites producing frameshifting mutations, a small exonic deletion missed by WES, and a novel deep intronic variant of unknown function. Thus non-coding variation (outside exons and flanking splice sites) accounted for only two likely disease-related variants among 400 GS alleles, with 4 other alleles unexplained after WGS. Among patients with isolated hypomagnesemia, six had monoallelic deletions in *HNF1B*. Additionally, a novel homoplasmic mitochondrial isoleucine tRNA variant was associated with hypomagnesemia and depression/anxiety in seven relatives spanning three generations who shared a maternal lineage. The electrolyte profiles of probands with disease-associated genotypes were consistent with the paradigmatic features associated with mutation in each gene.

## Introduction

Over the last decade, the frequency of non-coding variants with large effect sizes has been the subject of significant debate. The protein-coding coding genome comprises ∼1.5% of the human genome, with the non-coding genome accounting for the remainder. Non-coding regions comprise both transcribed elements, such as long non-coding RNAs and miRNAs, regulatory segments including enhancers and promoters, and highly repetitive sequences, often derived from transposable DNA elements^1^. Studies of Mendelian disorders, which are caused by variants with large effect size, are highly biased to coding regions and exon-flanking splice sites. As of March 24^th^ 2025, mutations in 4,970 genes have been linked to 1 or more Mendelian disease phenotypes^2^. In contrast, the number of loci in which rare non-coding mutations are the sole or predominant cause of large phenotypic effects is exceedingly small, with a few dozen examples, such as mutations in distal enhancer elements that regulate IHH expression in patients with craniosynostosis, mutations in the promoter region of the gamma-globin genes linked to hereditary persistence of fetal hemoglobinemia, and mutations in long non-coding RNAs linked to neurodevelopmental disorders ^3-5^.

In contrast, common variants with much smaller effect sizes identified by genome-wide association studies (GWAS) are vastly more frequent in non-coding regions, where they typically change the risk of a trait by <20%, predominantly by modulating gene expression.^6,7^ It has been suggested that the paucity of non-coding mutations with large effect is attributable to ascertainment bias, since the cost of whole genome sequencing has until recently been so expensive, however Mendelian loci identified from initial unbiased localization of genes by analysis of linkage in families have very rarely led to identification of non-coding variation as the sole or primary cause. Nonetheless, some probands with well-characterized Mendelian disorders predominantly caused by coding variants remain unsolved by WES, suggesting that these cases might be caused by non-coding variants. As the cost of WGS has come down dramatically, efforts to identify rare mutations with large effect have become more feasible.^8,9,10,11,12^.

We have investigated the spectrum of mutations underlying known Mendelian diseases by the study of patients with renal electrolyte disorders. These include hypokalemia, typically caused by salt wasting, of which the most frequent genetic cause is Gitelman syndrome (GS), a recessive disorder affecting about 1/40,000 people. GS results from biallelic LoF variants in the gene *SLC12A3* encoding the thiazide-sensitive electroneutral Na-Cl cotransporter (NCC), of the distal convoluted tubule (DCT). The loss of salt reabsorption in the DCT leads to salt wasting, reduced blood pressure, and activation of the renin-angiotensin system, which increases electrogenic Na+ reabsorption in the distal nephron, leading to increased excretion of K+ and H+, producing hypokalemia and alkalosis. Altered DCT salt reabsorption also results in hypomagnesemia and hypocalciuria. GS is typically diagnosed in the second decade of life. In large pedigrees, penetrance of biallelic GS genotypes is essentially 100% while monoallelic mutations cause much milder phenotypic expression.^13,14^

Mutations in genes that impair salt reabsorption in the thick ascending limb of Henle (*NKCC2, KCNJ1, CLCNKB, BSND, MAGED2*) cause Bartter syndrome, which is distinguished by severe salt wasting, prenatal or neonatal presentation, and hypercalciuria.^15-17^ Recessive loss of Na+ reabsorption in the distal nephron by genes encoding the three subunits of ENaC, the electrogenic Na+ channel of the distal nephron, causes pseudohypoaldosteronism type 1 featuring severe salt wasting, hyperkalemia, and metabolic acidosis, while monoallelic loss of the mineralocorticoid receptor that regulates ENaC activity causes a less severe PHA1 phenotype. Renal hypomagesemia can present with or without alkalosis and with hypo or hypercalciuria; mutations causing hypomagnesemia include biallelic mutations in *CLDN16, CLDN19, TRPV6, HNF1B* and mitochondrial mutations in gene encoding the isoleucine tRNA. The spectrum of phenotypes resulting from mutations that produce these phenotypes is shown in (**Table 1**).

**Table 1.**
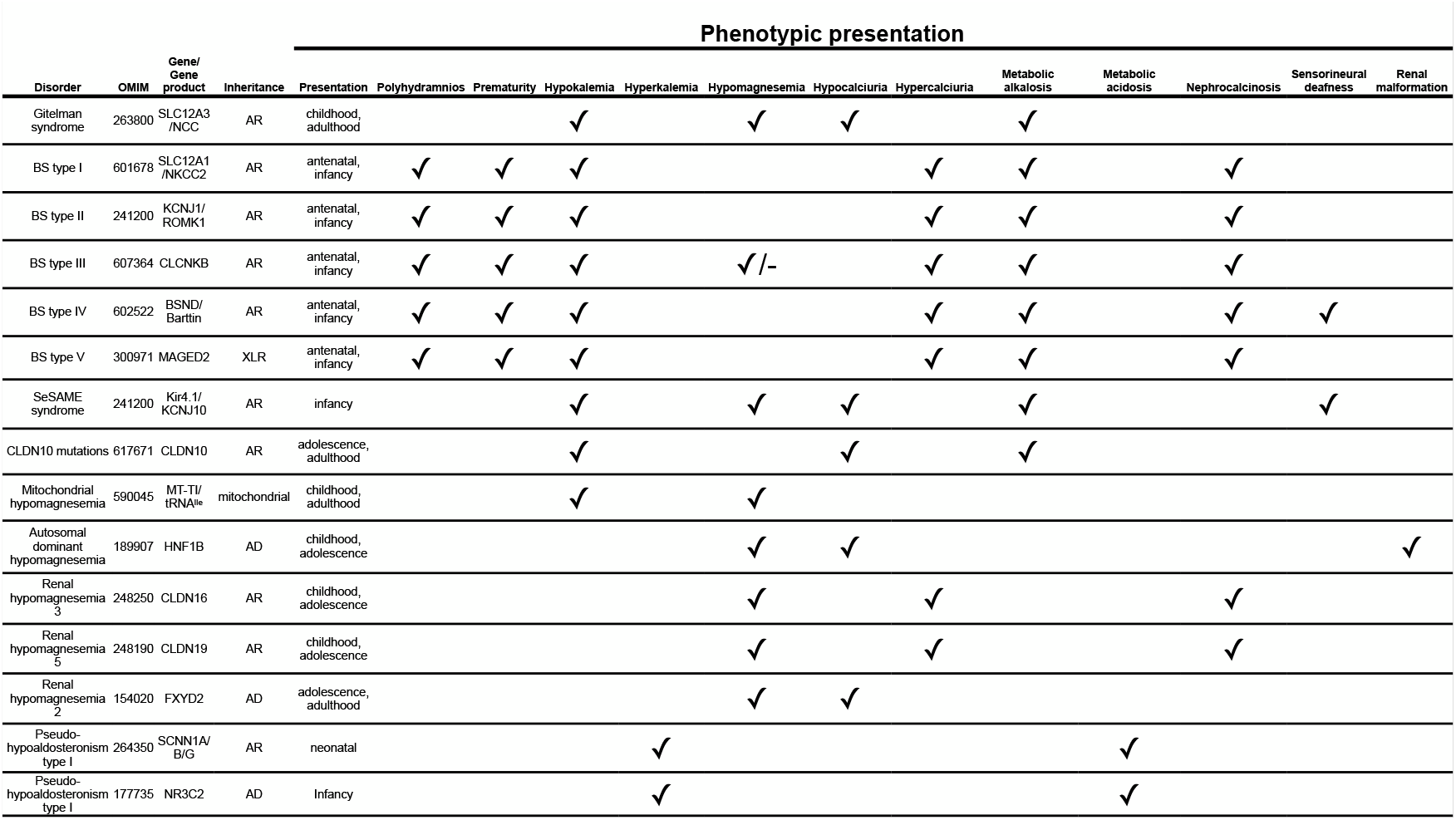
Mendelian forms of salt-wasting disorders. Mendelian salt-wasting disorders are organized by phenotypic features.

Over many years, our laboratory received samples from patients with suspected salt wasting and/or renal electrolyte disorders referred by physicians. The degree of clinical study was variable. To better understand the contribution of coding and noncoding variants to GS and other renal electrolyte disorders, whole exome sequencing of peripheral blood DNA was performed on 329 unrelated patients with a documented or suspected renal salt-wasting disorder. We report herein the results of these studies.

## Methods

### Cohort characteristics and DNA sequencing

329 patients with suspected salt-wasting disorders were referred for genetic analyses. The Yale IRB approved the study (approval # 9512008556). Previously, 108 members of this cohort were described and identified as having biallelic mutations in *SLC12A3*.^14^ The sample IDs used in this study were not known to anyone outside the research group.

The available clinical features of the remaining 221 subjects are shown in **Table S1**. These included 221 patients with hypokalemia (serum K^+^ < 3.5 mM), 172 of whom were taking K^+^ replacement; 196 with hypomagnesemia (serum Mg^2+^ < 1.8 mM), 106 taking Mg^2+^ replacement, and 103 with hypocalciuria (urinary Ca^2+^: creatinine ratio < 0.2 [mmol/mmol]). Thirty-four patients had no available clinical data other than reported salt wasting. Summary electrolyte values within the cohort is shown in (**Fig. S1)**.

DNA from the 222 probands not previously analyzed was extracted from venous blood samples or saliva and subjected to WES for genetic analyses at the Yale Center for Genome Analysis (YCGA); summary sequencing statistics are in **Table S2**.

Variants across all genes were called if they had ≥ 8 independent reads, at least 3 independent variant calls, and a genotype quality (GQ) ≥ 20. Copy number variants were identified using GATK’s germline copy number variant tool^18^. They were annotated using ANNOVAR^19^ for allele frequency in the gnomAD and BRAVO databases. Likely damaging variants were defined as those with allele frequency < 0.001 in gnomAD and BRAVO and were LoF (nonsense, canonical splice-site, frameshift indels, and start loss) and or classified as damaging missense by CADD score ≥ 20 ^20^ and/or MetaSVM-D ^21,22^, and/or classified as pathogenic or likely pathogenic by ClinVar. Variants of interest that passed these filters were visually confirmed.

### Gene burden analysis

All damaging biallelic genotypes identified through whole exome sequencing were statistically analyzed using a one-tailed binomial test to test for departure from the expected distribution of biallelic genotypes for damaging variants in all genes in patients. This method compares the observed rare damaging recessive genotype variants within a gene to the expected mutation frequency due to a gene’s intrinsic mutability. The expected mutation frequency was determined using a polynomial regression model coupled with a one-tailed binomial test derived from the de novo mutation rate of each gene in the genome within a cohort of 2,871 probands with congenital heart disease and 1,789 unaffected controls^23^.

### Whole genome sequencing and analysis

Whole genome sequencing of selected samples was performed to a mean depth of 37 indpendent reads per base pair by ligating adapters with appropriate dual multiplexing indices to the ends of target DNA fragments, followed by hybridization to Illumina flow cells and cluster generation, followed by 151 base paired-end sequencing reads following Illumina protocols on the NovaSeq 6000. Samples were subjected to 10bp indexing reads following the completion of read 1. 98.55% of all bases had at least 10 independent reads, sufficient to make confident calls of heterozygous variants in the vast majority of cases (**Table S3)**. A MAF<0.001 in gnomAD WGS was used as a threshold for variant filtering.

Ensembl Variant Effect Predictor (VEP) tool was used to annotate noncoding variants in noncanonical splice regions, promoter regions, 3’ UTR sites, and some transcription factor binding sites^24^. Large intragenic deletions were identified using both the GATK g-CNV pipeline and LUMPY^18,25,^. Deep intronic mutations that introduce putative novel splice sites were identified using SpliceAI, a tool that uses a deep neural network to predict cryptic loss of function from SNVs^26^. Variants of interest that passed these filters were visually confirmed.

## Results

The demographic features of the cohort are shown in (**Table 2**).^27^ A Q-Q plot comparing the observed and expected frequency of damaging biallelic variants for all genes by the binomial test (see Methods) is shown in **Fig. 1**. The burden of damaging recessive genotypes was highest for the Gitelman syndrome gene *SLC12A3* (197 recessive genotypes) followed by the Bartter syndrome genes *CLCNKB* (n = 36), *SLC12A1* (n = 13), *KCNJ1* (n = 7) and *BSND* (n =2). Two males had damaging hemizygous mutations in the X-linked gene *MAGED2*. There were also 6 heterozygous deletions removing exons of the known dominant hypomagnesemia gene *HNF1B*, and one proband with hypomagnesemia had a novel homoplasmic mutation in the mitochondrial tRNA^ile^ (**Fig.1**). Lastly, two patients had damaging recessive genotypes in *SLC26A3*, which encodes the colonic Na^+^/HCO_3_^-^ exchanger whose mutation causes salt loss.

**Table 2.** Salt-wasting proband demographic information. Ethnicity was determined by principal component analysis compared to HapMap samples using EIGENSTRAT.

|  |  |
| --- | --- |
| **Sample size** | 329 |
| **Gender** |  |
| Male | 152 (46%) |
| Female | 177 (54%) |
| **Ethnicity** |  |
| European | 274 (83%) |
| Mexican | 18 (6%) |
| African | 17 (5%) |
| East Asian | 10 (3%) |
| South Asian | 10 (3%) |

**Fig. 1.**
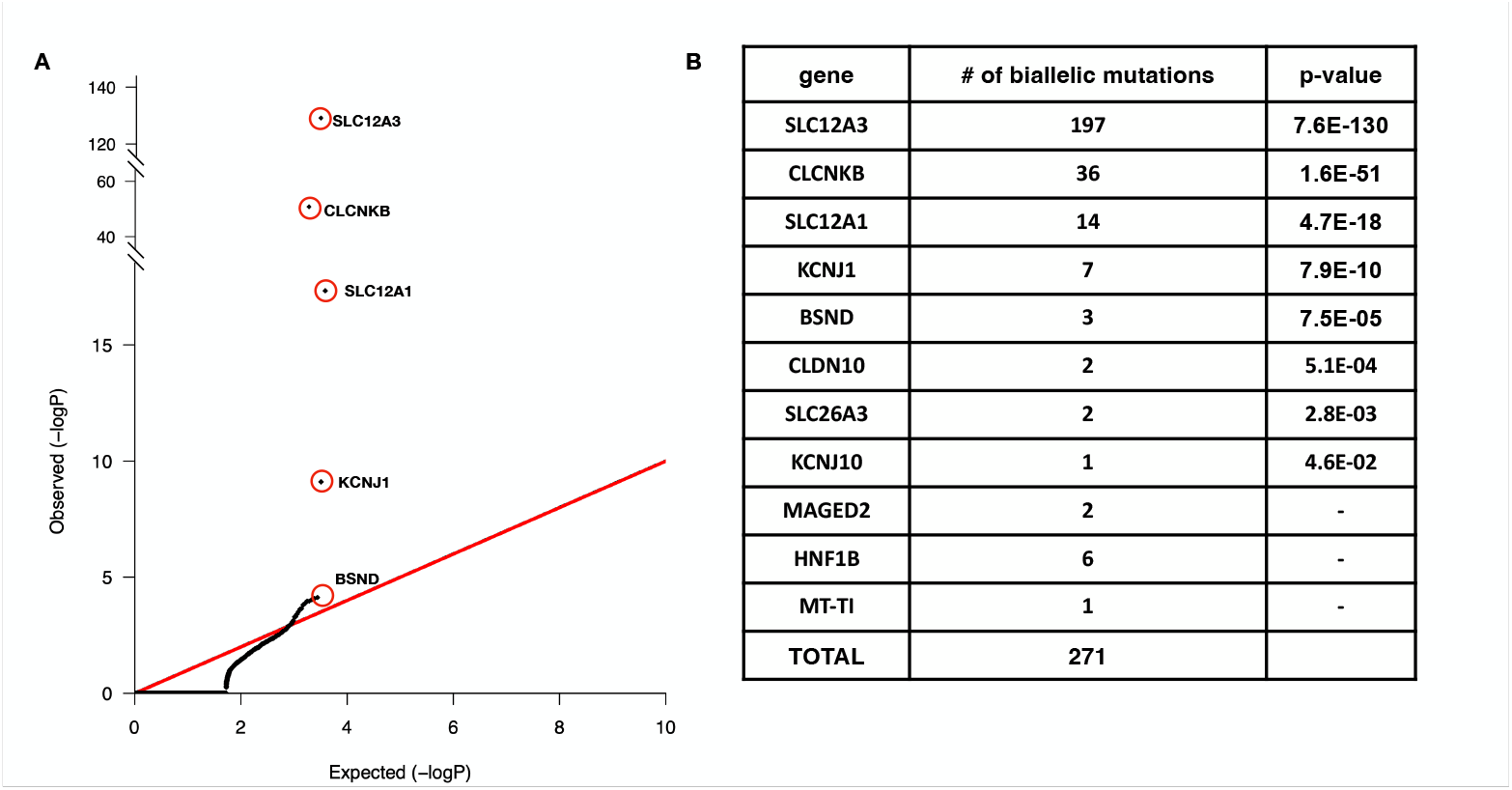
Binomial test on recessive genotypes in the salt-wasting cohort. (A) Recessive genotypes shown include LOF, D-mis, and non-frameshift insertions or deletions. The expected number of RGs in each gene was calculated from the total number of observed RGs described in the Methods. The significance of the difference between the observed and expected number of RGs was calculated using a one-sided binomial test. While the observed values closely conform to expected values in controls, SLC12A3, CLCNKB, and SLC12A1 show a significantly increased burden of RGs in cases. (B) Results from binomial analysis on 329 salt-wasting probands. Top 8 genes from the binomial test are listed as well MAGED2, HFN1B and MT-TI who represent 2 individuals with hemizygous damaging mutations, 6 individuals with heterozygous deletions as well as a homoplasmic mitochondrial mutation, respectively. Genes with genome-wide significant enrichment are in bold (P-value < 2.6 × 10-6 [0.05/19347]).

The observed biochemical features and age of diagnosis in probands with presumed disease-causing genotypes in each gene closely matched the expected features (**Fig. 2)**. Patients with *SLC12A3, CLCNKB* and BSND mutations had lower K+ levels than those with SLC12A1 or KCNJ1 mutations, and patients with *MAGED2* mutation had hyperkalemia rather than hypokalemia as expected. Patients with *SLC12A3* mutations and had significant hypomagnesemia, as did the patient with mitochondrial mutation and HNF1B mutations, but others did not. As expected, Gitelman and CLDN10-mutant patients had hypocalciuria, while Bartter patients had hyper- or normal calciuria. Patients with Gitelman, *CLDN10* and the mitochondrial mutation patients were predominantly diagnosed in adolescence or adulthood, while Bartter and MAGED2 patients were diagnosed in the first years of life. In sum, the consistency of these phenotypes provides compelling evidence that these inferred disease genotypes are not false-positive, and that there is little variability in phenotypic expression in patients with mutations in these genes.

**Fig. 2.**
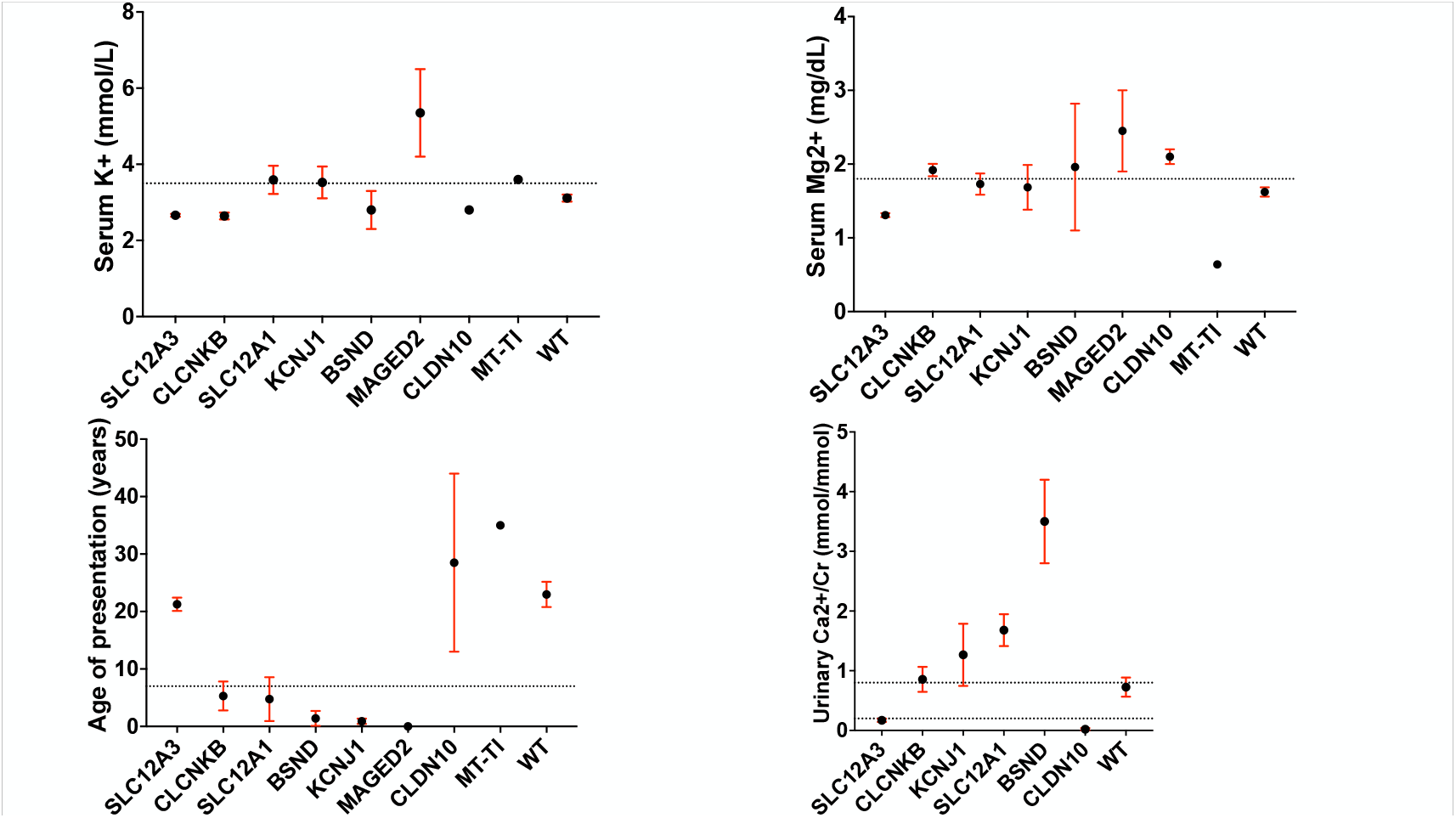
Serum K+ values for biallelic mutations in genes linked to Mendelian forms of salt wasting. Each point indicates the mean for each sample group or the value for an individual patient. Dashed lines indicate normal value cutoffs. Errors bars indicate standard error.

The 394 rare damaging alleles in the 197 probands with biallelic rare damaging mutations in *SLC12A3* included of 140 LoF alleles and 254 damaging missense alleles. There were 46 homozygous and 151 compound heterozygous genotypes. There were 139 different variants, including 92 found once and 47 that were recurrent. The most frequent variant was the G741R mutation, which was found 37 times, representing 9.9% of the mutant alleles. This variant, is enriched 316-fold compared to its frequency of 3 × 10^−4^ in the gnomAD population. Two canonical 5’ donor splice site variants, c.1180+1G>T and C.2883+1G>T, together accounted for 57 mutant alleles. The mean/median frequency of all mutant *SLC12A3* alleles in the gnomAD database was 1.5 × 10^−4^, and 46 variants had an allele frequency of zero among >200,000 alleles in gnomAD; 44 had not been previously reported as Gitelman-related and were identified in 54 patients with a recessive genotype. Of these, 44 had characteristic biochemical findings of hypokalemia and either hypomagnesemia, or hypocalciuria. A complete list of all biallelic genotypes in *SLC12A3* found in these probands can be found in (**Table S4** and **Table S5**).

Damaging alleles included three unusual mutations at splice donor and acceptor sites predicted to abolish gene function: a previously unreported 6 bp duplication at the intron 15 splice acceptor site that is expected to duplicate a serine-proline dipeptide in the encoded protein.

There were six intragenic deletions spanning two or more exons; three deletions were previously reported, while three were novel, including one spanning exons 1-7 and another found in siblings.

### Mutations at non-GT/AG splice donor and acceptor sites in *SLC12A3*

Three previously unreported noncanonical splice variants were identified that result in the disruption of wild-type splice donor or acceptor sites. The three mutations identified were predicted to result in loss of function using SpliceAI. These intronic variants disrupt normal splicing and are predicted to be pathogenic, leading to exon skipping, frameshifts, and protein truncation.

One variant in *SLC12A3* splicing is found in a GS proband with hypokalemia, hypomagnesemia and a pathogenic LoF monoallelic variant. WES revealed that this patient also has a previously unreported non-coding variant, 16-56947166 T>A (c.2952-10T>A), 10 bp upstream of exon 26 (**Fig. S2A**). This mutation lies in the conserved polypyrimidine tract upstream of the canonical AG acceptor sequence and creates a new ‘AG’ site that Splice AI predicts to be a strong splice site. In parallel, the T>A mutation would further degrade the already weak WT polypyrimidine tract at the end of intron 25, making it less favorable (**Fig. S2B, Fig. S2C**)^28,29,30^. Use of this new splice site is predicted to introduce a frameshift insertion into the encoded protein, resulting in premature termination, deleting the last 46 amino acids (**Fig. S2D**). In addition to this region being highly conserved among vertebrates, this deletion is highly likely to be pathogenic given that a pathogenic variant that only removes the last 12 residues of SLC12A3 has been previously identified in Gitelman syndrome patients^31^.

The second novel variant alters splicing by disrupting the splice donor site at the exon 25/intron 25 junction, introducing a G>A mutation at the 5^th^ base of the intron, altering a ‘G’ that is nearly invariant among splice donor sites. (**Fig. S3A**). The position of this variant is also extremely well conserved among vertebrate orthologs, consistent with this site being intolerant to mutation (**Fig. S3B**). Moreover, SpliceAI predicts with high confidence (score of 0.88, high precision cutoff >0.8) that this variant will result in loss of splice donor function at this location (**Fig. S3C**), leading to exon skipping, and producing a deletion and frameshift, producing a truncated version of SLC12A3 that is missing its last 46 residues (**Fig. S3D, E**).

The third novel splice variant occurs 2 bp downstream of the “GT” splice donor site following exon 16, altering ‘A’, which is highly conserved among vertebrate orthologs, to ‘G’ (**Fig. S4A**), and falls within the conserved extended splice donor site sequence ‘**GT**RAGT’ (**Fig. S4B**).^32^ SpliceAI predicts that this variant results in splice donor loss (**Fig. S4C**). The splice-disrupting variant results in a frameshift that effectively removes nearly half of the full-length protein and would result in loss of function (**Fig. S4D**).

In addition, WES analysis of GIT614-1 identified a novel heterozygous 6 bp duplication spanning the end of intron 15 and the first three base pairs of exon 16; this alters the wild-type 5’-**CAG**CCC-3’ sequence to 5’-**CAG**CCC**CAG**CCC-3’ (**Fig. 3A**). The proximal CAG in the mutant allele is strongly favored (> 99% probability by NNSPLICE which handles larger indels^33^). The spliced mRNA is thus predicted to contain 6 additional bases, adding a serine-proline dipeptide to the wildtype sequence at the start of exon 16 before the proline-glutamine at positions 642 and 643 found in wildtype (**Fig. 3B,C**). This mutation has not to our knowledge been reported previously.

**Fig. 3.**
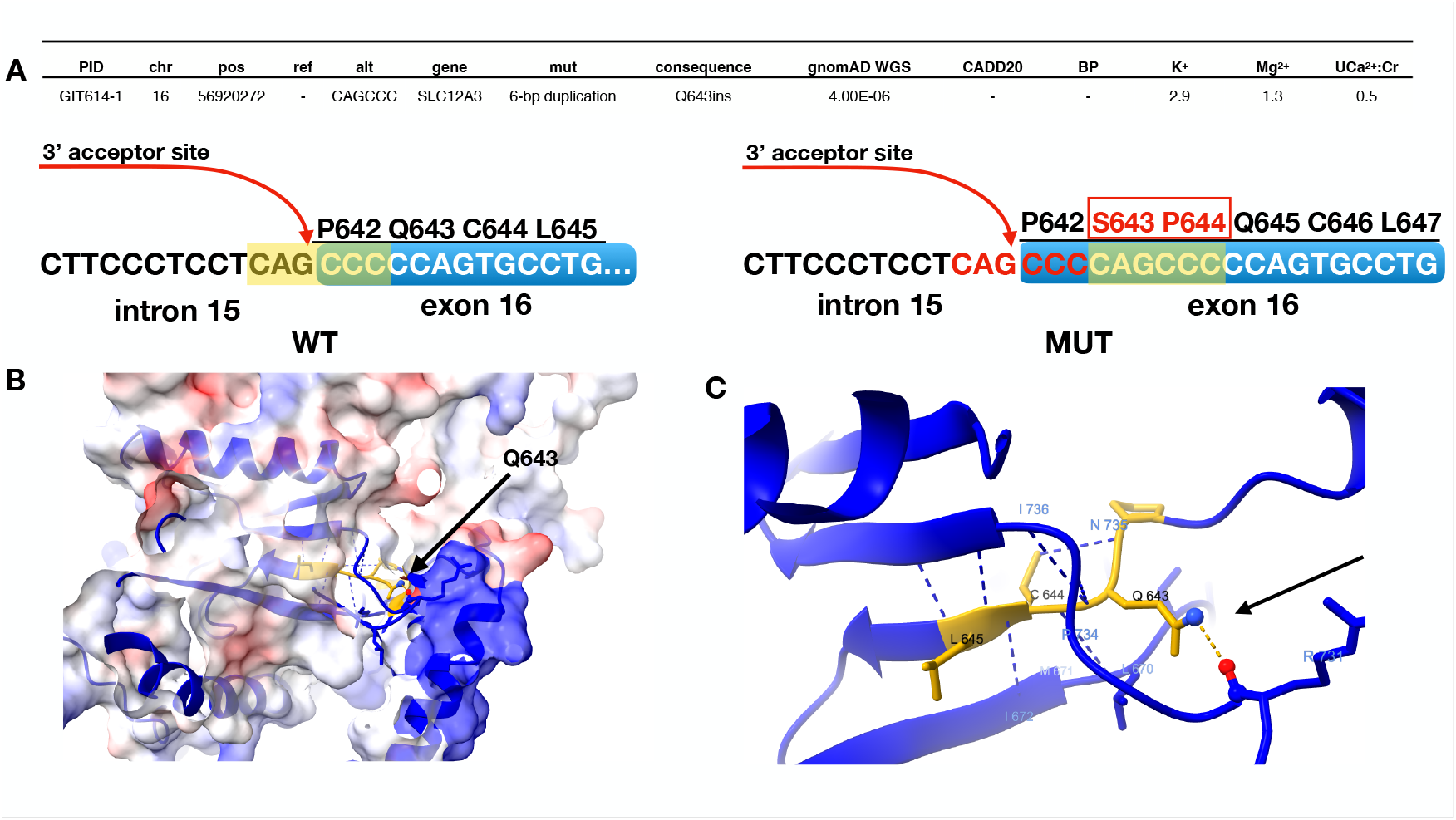
6-bp duplication of splice acceptor site at exon 16 3’ results in cryptic insertion of the dipeptide. (A) Information on 6-bp insertion at 3’ acceptor site followed by clinical features of GS proband. Side-by-side comparison of wild-type exon 16 splice acceptor site and cryptic 3’ acceptor site 6 bp downstream of the true acceptor site. The 6-bp insertion on the 5’ end of exon 16 results in the insertion of serine and proline at Q643. (B) Q643 lies within a deep pocket of the cytoplasmic tail of SLC12A3. (C) The amine group of Q643 is predicted to hydrogen bond to the carbonyl group of R731. The resultant Q643S from the 6 bp insertion would be expected to disrupt the dipole-dipole interaction.

### Identification of small intragenic deletions of two or more exons in *SLC12A3*

GATK gCNV was used to identify putative structural variants within *SLC12A3* using WES data. A total of six intragenic deletions were identified by this analysis. Five out of six of these GS probands had hypokalemia and hypomagnesemia, with the additional finding of hypocalciuria in one proband in whom urinary calcium was measured (**Fig. 4A**). These patients had identified monoallelic mutations in each of these six probands, including 3 ClinVar pathogenic D-mis mutations and 3 LoF variants. Three of these deletions are previously reported as pathogenic; one of these deleted exons 2 and 3 in a proband; another deleted exons 4-6 in an additional two probands, all identified by GATK gCNV (**Fig. 4B**). The 5’ and 3’ boundaries of these deletions had a >90% sequence match to that of Alu elements, consistent with occurrence via recombination between these sequences (**Fig. 4B**). These intragenic deletions were confirmed using IGV, which showed a ∼50% reduction in the depth of independent read coverage across the deleted interval in all affected patients (**Fig. S5** and **Fig. S6**).

**Fig. 4.**
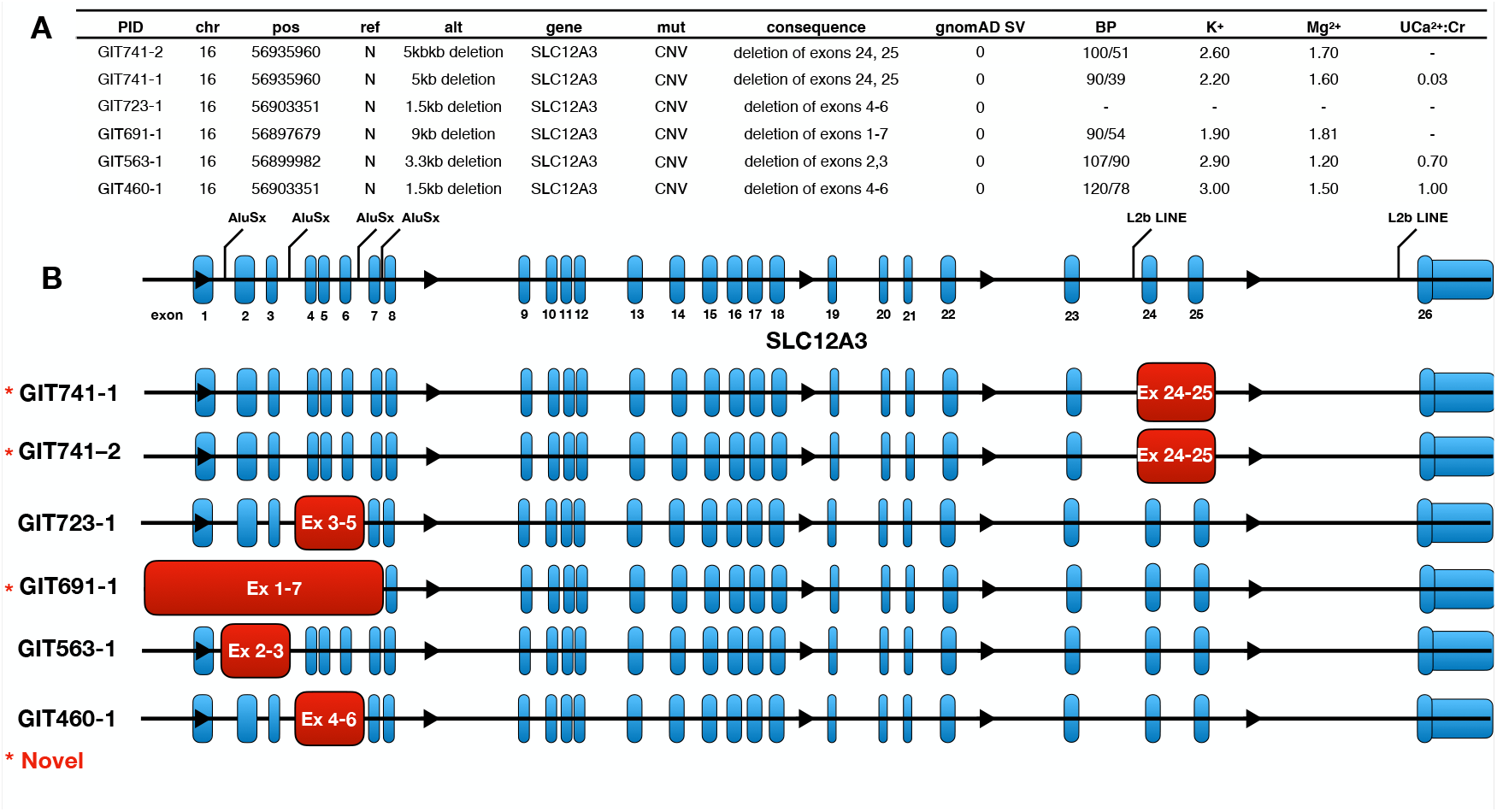
List and diagram of intragenic deletions of 2 or more exons in SLC12A3. (A) List of intragenic structural variants in 6 salt-wasting probands followed by blood pressure, electrolyte data, and urinary Ca2+:Cr ratio. (B) Schematic showing the location of the 6 intragenic deletions (red) in SLC12A3 (exons in blue, noncoding regions in black) and Alu sites that flank the 5’ and 3’ ends of the deletions.

There were also three previously unreported monoallelic deletions in *SLC12A3*. Two of these span exons 24 and 25, and the other removes exons 1-7, including the promoter region and start codon. Interestingly, the two probands that carry the exon 24 and 25 deletions are a set of siblings. This was confirmed using an identify-by-descent pairwise calculation in PLINK, which resulted in an estimated 48.5% sharing of DNA sequence identity^34^. Although a LINE and Alu element was found at one of the borders of the deletions, homology with this sequence was not identified at the other end of the deletion in the wildtype DNA sequence (**Fig. S7A**). Examination of the sequence alignment across SLC12A3 on IGV confirmed that both siblings carry the same deletion (**Fig. S7B**). Non-homologous end joining of a double strand break seems a plausible explanation for the genesis of this deletion.

A previously unreported 9 kb deletion that begins 2 kb upstream of the *SLC12A3* promoter sequence and ends within intron 7 was identified in another GS proband with a monoallelic variant. This deletion contains a single Alu element at its 3’ end (**Fig. S8A**). Although this deletion can be visualized on IGV (**Fig. S8B**), we sought other methods to validate the presence of this mutation given that it was present in a single proband. We sought support for this deletion by loss of heterozygosity in the 9 kb segment (**Fig. S9**). Among 68 SNPs with minor allele frequency ≥ 20%, we observed no heterozygous SNPs in the deleted interval inferred by sequencing, while flanking sequence showed typical patterns of heterozygosity, strongly supporting evidence of this deletion.

### Whole-genome analysis of 7 probands with monoallelic mutations IN *SLC12A3*

The estimated prevalence of GS of 1/40,000 implies a total allele frequency of ∼1/200. After excluding the 271 probands in the current cohort with a likely identified genetic cause, the expected number of individuals with a single *SLC12A3* variant among the remaining 59 subjects was ∼0.6. We instead identified 7 such subjects, far more than expected by chance. The single variants in these individuals included 3 rare LoF, and 4 rare ClinVar-pathogenic missense variants (**Table 3**). These subjects had available electrolyte levels, and mean serum K^+^ was 2.56mM, mean Mg^2+^ was 1.34 mg/dl, and mean HCO_3_^-^ was 29 mM, not different from values seen in patients with biallelic mutations in *SLC12A3* and distinct from those found in wild-type subjects and those with known heterozygous variants (**Fig. S10**).^13^ No damaging mutations in other known renal electrolyte genes were identified in these subjects, providing no support for digenic contributions to disease.

**Table 3.**
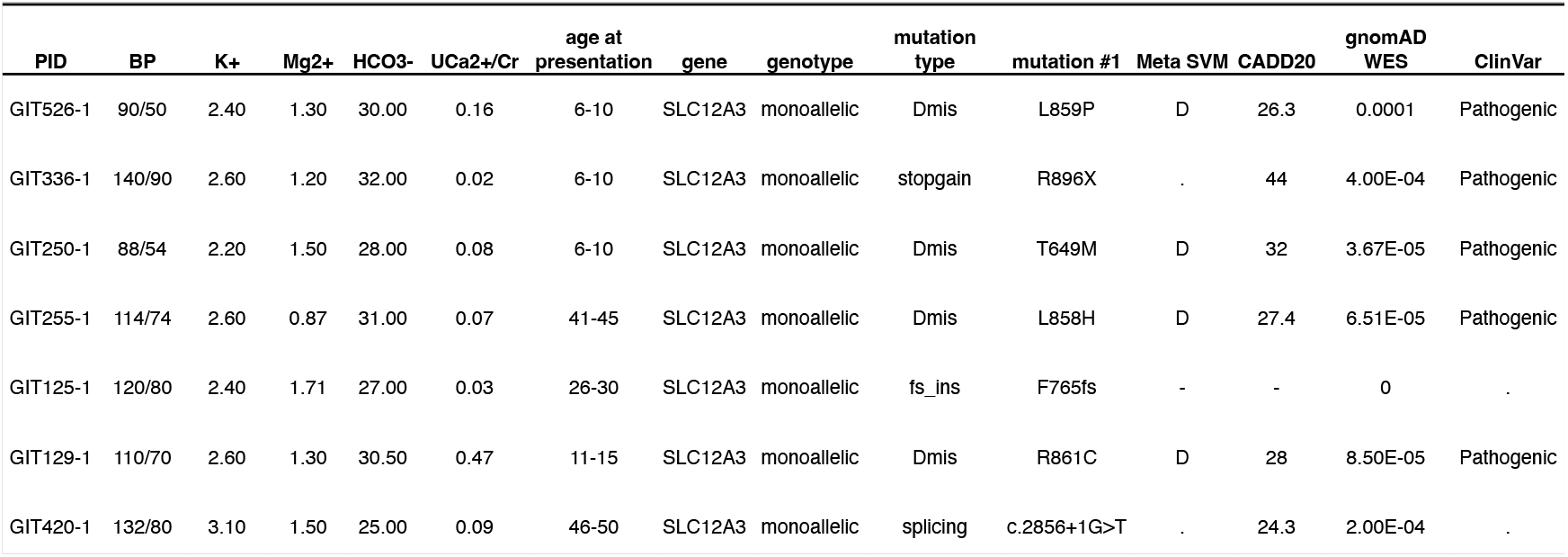
List of 7 probands with Gitelman syndrome that carry damaging monoallelic mutations in SLC12A3. Clinical information, followed by information on the monoallelic mutations is shown.

| PID | BP | K+ | Mg2+ | HCO3- | UCa2+/Cr | age at presentation | gene | genotype | mutation type | mutation #1 | Meta SVM | CADD20 | gnomAD WES | ClinVar |
| --- | --- | --- | --- | --- | --- | --- | --- | --- | --- | --- | --- | --- | --- | --- |
| GIT526-1 | 90/50 | 2.40 | 1.30 | 30.00 | 0.16 | 6-10 | SLC12A3 | monoallelic | Dmis | L859P | D | 26.3 | 0.0001 | Pathogenic |
| GIT336-1 | 140/90 | 2.60 | 1.20 | 32.00 | 0.02 | 6-10 | SLC12A3 | monoallelic | stopgain | R896X | . | 44 | 4.00E-04 | Pathogenic |
| GIT250-1 | 88/54 | 2.20 | 1.50 | 28.00 | 0.08 | 6-10 | SLC12A3 | monoallelic | Dmis | T649M | D | 32 | 3.67E-05 | Pathogenic |
| GIT255-1 | 114/74 | 2.60 | 0.87 | 31.00 | 0.07 | 41-45 | SLC12A3 | monoallelic | Dmis | L858H | D | 27.4 | 6.51E-05 | Pathogenic |
| GIT125-1 | 120/80 | 2.40 | 1.71 | 27.00 | 0.03 | 26-30 | SLC12A3 | monoallelic | fs_ins | F765fs | - | - | 0 | . |
| GIT129-1 | 110/70 | 2.60 | 1.30 | 30.50 | 0.47 | 11-15 | SLC12A3 | monoallelic | Dmis | R861C | D | 28 | 8.50E-05 | Pathogenic |
| GIT420-1 | 132/80 | 3.10 | 1.50 | 25.00 | 0.09 | 46-50 | SLC12A3 | monoallelic | splicing | c.2856+1G>T | . | 24.3 | 2.00E-04 | . |

We considered it likely that these subjects had second SLC12A3 mutations that were not detected by WES. To search for these mutations, DNA from these 7 individuals was subjected to whole genome sequencing and analysis (**Fig. S11**). The range of potential noncoding variants with large effect include deep intronic mutations that result in a gain of splice sites leading to altered protein sequence, intragenic deletions of one or more exons not detected by WES, and mutations in putative regulatory sites such as enhancers and promoters.

Among these 7 monoallelic probands, 4 rare non-coding variants in *SLC12A3* were identified by WGS, 3 of which are likely pathogenic.

### Deep intronic variants that disrupt splicing

In patients GIT250-1 and GIT255-1, who have different known pathogenic missense variants in *SLC12A3*, an identical deep intronic variant, 16-56917770 C>T, was identified (**Fig. S12A**). SpliceAI determined that this deep intronic variant introduced a probable GT splice donor site deep within intron 13 of SLC12A3, with a probability score of 0.72, well above the webtools ‘recommended’ cutoff of 0.5 for splicing variant candidates (**Fig. S12B**). This variant has a prevalence of 3 × 10^−5^ in gnomAD. Most interestingly, the identical homozygous mutation was previously identified in a Taiwanese GS cohort. The functional significance of this mutation was shown in the prior paper by sequencing cDNA from kidney tissue samples from these patients, which showed a 238 bp insertion between exons 13 and 14 resulting from insertion of a cryptic exon via the new splice donor site created by the point mutation.^35^ Interestingly, one of the current probands is of East Asian descent while the other is of Mexican ancestry, suggesting the latter mutation may have arisen independently.

### Identification of a small intragenic deletion of two or more exons in *SLC12A3*

LUMPY was used to identify a structural variant within SLC12A3 that was not detected by WES analysis. An intragenic deletion was identified in a GS proband that removes exons 2 and 3 that had hypokalemia, hypomagnesemia, and hypocalciuria (**Fig. S13A**). Whole exome sequencing had identified a monoallelic ClinVar pathogenic D-mis mutation in this proband (**Fig. S13B**). Interestingly, this exact deletion was detected in another proband using GATK gCNV on WES data but was not identified in this subject.

### Rare intronic variant of unknown function

Proband GIT336-1 presented with clinical features that are pathognomonic for Gitelman syndrome and was found to carry a heterozygous premature termination codon, R896X, that removes 125 amino acids from the c-terminal end of SLC12A3. WGS identified a noncoding variant at position 16-56900664 (c.283-318 G>A) deep within intron 1 (**Fig. S14A**). Parental sequences showed that this variant was in trans to the LoF variant (**Fig. S14B**,**C**). This variant has not been reported in the gnomAD WGS database. SpliceAI and gatkCNV failed to identify any intronic mutations that disrupt splicing or any exonic deletions in *SLC12A3*. This variant is at a position well conserved among vertebrates and invariate among primates (**Fig. S14D**), and is located ∼1.5kb downstream of the *SLC12A3* transcription start site. This variant is not known to lie in an enhancer, and whether it contributes to pathogenesis is presently unknown.

WGS did not identify a compelling noncoding mutation in three GS probands that had monoallelic likely pathogenic variants, and all had pathognomonic features of GS (**Table S6**). Searches included examination of likely splice branch points and exonic splice enhancers.

Collectively, among the total of 404 likely pathogenic variants in *SLC12A3*, there were only 3 that were not identified by WES, and only two that did not alter coding regions or were within the extended splice recognition sequences.

### Maternally inherited hypomagnesemia caused by homoplasmic mitochondrial mutation

We previously identified a pathogenic homoplasmic mutation, m. 4191 U>C, in the mitochondrial isoleucine tRNA linked to maternally inherited hypomagnesemia in 142 members of one family^36^. Given the prospect of identifying mitochondrial mutations in unexplained cases of hypomagnesemia, we directed our genetic analyses to the mitochondrial genome, which is sequenced in the process of WGS and annotated by GATK and ANNOVAR.

Proband GIT783-1 was referred for genetic analyses due to unexplained hypomagnesemia. Her symptoms initially arose during pregnancy in her early 20s. She presented in her 30s with weakness and fatigue and was found to have an extremely low magnesium level of 0.98 mg/dL and a fractional excretion of Mg^2+^ of 11.4%, which is inappropriately high. Other electrolytes were normal (**Fig. 5A**). She did not tolerate oral magnesium due to intractable diarrhea or IV replacement of magnesium, which required administration over 12 hours each day, which produced palpitations, lightheadness, and flushing. She had an initial trial of peritoneal magnesium infusions but these were not tolerated due to abdominal pressure and cramping.^37^ She eventually tolerated a decreased concentration of 2g MgSO4 in 500 ml split over two daily infusions, which has kept her magnesium levels around 1.5 mg/dL. She also suffers from major depressive disorder, arrhythmias, paresthesias, syncope, nocturia, and soft tooth enamel. A brain MRI identified no pathology. Whole exome analyses did not reveal biallelic mutations in any known Mendelian disorders of electrolyte homeostasis.

**Fig. 5.**
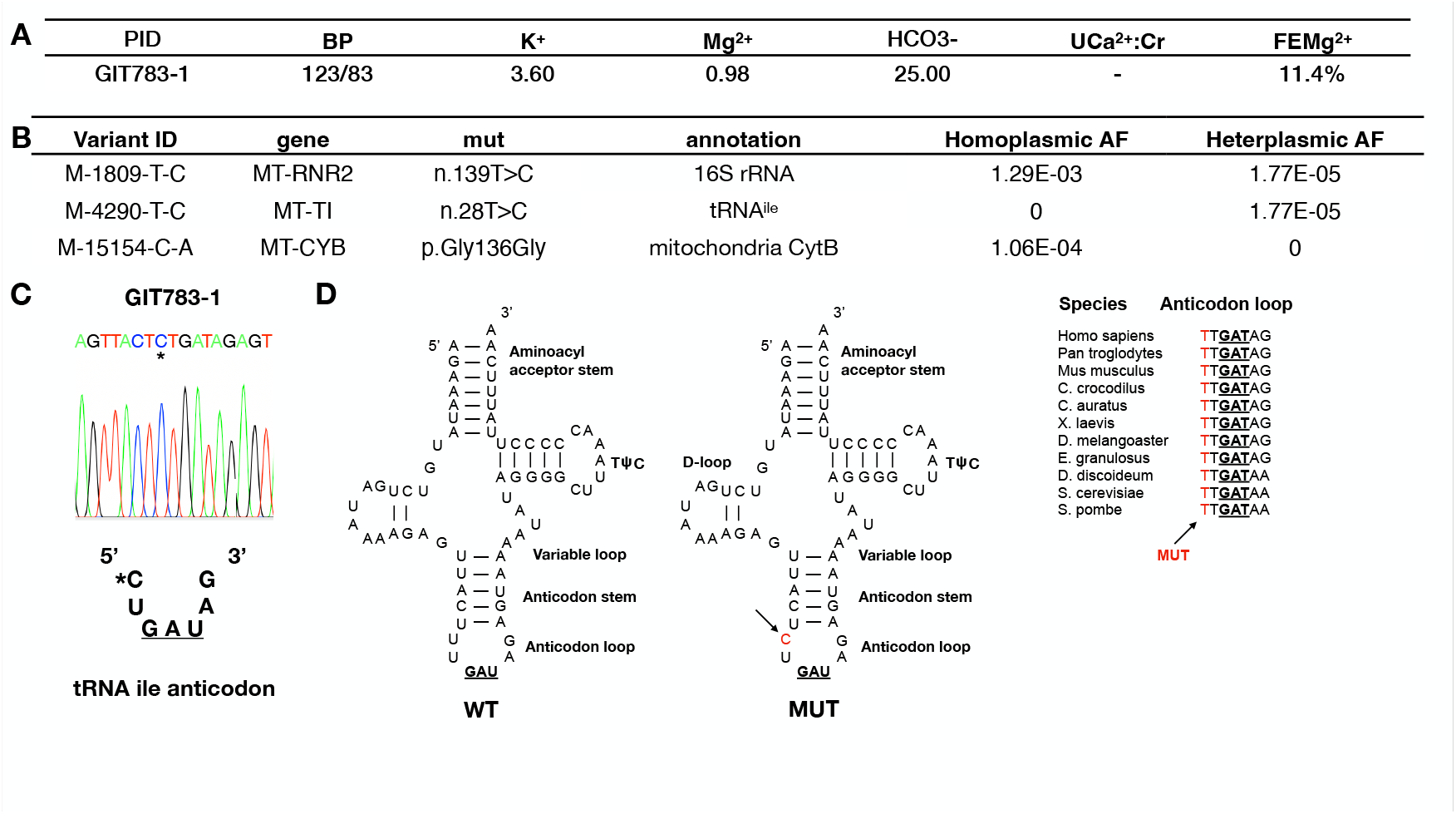
Homoplasmic variant in mitochondrial tRNA ile next to pre-identified mutation. (A) Clinical features of proband GIT783-1, which are listed as mg/dL. (B) List of three mutations that map to locations within the mitochondrial genome. Homoplasmic and heteroplasmic frequencies from gnomAD are shown. (C) Targeted sequencing showing mitochondrial mutation m.4291T>C identified in patients with Gitelman-like syndrome followed by m.4290T>C found in a kindred with maternally inherited hypomagnesemia. (D) Annotated secondary structure of tRNAIle with the anticodon highlighted in red and m.4290T>C in pink. Conservation of m.4290 within a select group of species is shown to the right.

Analyses of her mitochondrial genome revealed three homoplasmic variants. One of these results in a synonymous substitution in mitochondrial cytochrome B, another is a T>C substitution in MT-RNR2, the mitochondrially-encoded 16S rRNA. Both of these variants have been found as homoplasmic variants in gnomAD, not associated with disease. Remarkably, the third variant produced a T>C substitution at m.4290 in the mitochondrial tRNA^Ile^ (MT-TI) (**Fig. 5B**). This variant was homoplasmic and lies one base pair 5’ to the m.4291 T>C homoplasmic mutation previously shown to result in maternally-transmitted hypomagnesemia^35^ (**Fig. 5C**).

These two bases lie immediately 5’ to the anticodon of this tRNA; moreover, these two mutated positions are invariant from yeast to humans. (**Fig. 5D**).^38^

We recruited available family members of the proband, collected electrolyte data, and sequenced genomic DNA. The results showed that all six additional family members on the same maternal lineage as the proband had hypomagnesemia, with values ranging from 0.9 to 1.7 mg/dL; all five for whom a DNA sample was available had the homoplasmic m.4190 U>C mutation, and the 6^th^ subject is the deceased matriarch of the maternal lineage, who is an obligate carrier (**Fig. 6**). Moreover, one relative who was not on the maternal lineage was sampled and did not have the mutation and was normomagnesemic, making an environmental cause of hypomagesemia unlikely. The finding that all members of the family on the same maternal lineage had both the rare homoplasmic variant and hypomagnesemia, as well as the prior demonstration of the role of a nearly identical homoplasmic variant in hypomagenesemia, provides compelling evidence of the causal role of the variant in this family with hypomagnesemia.

**Fig. 6.**
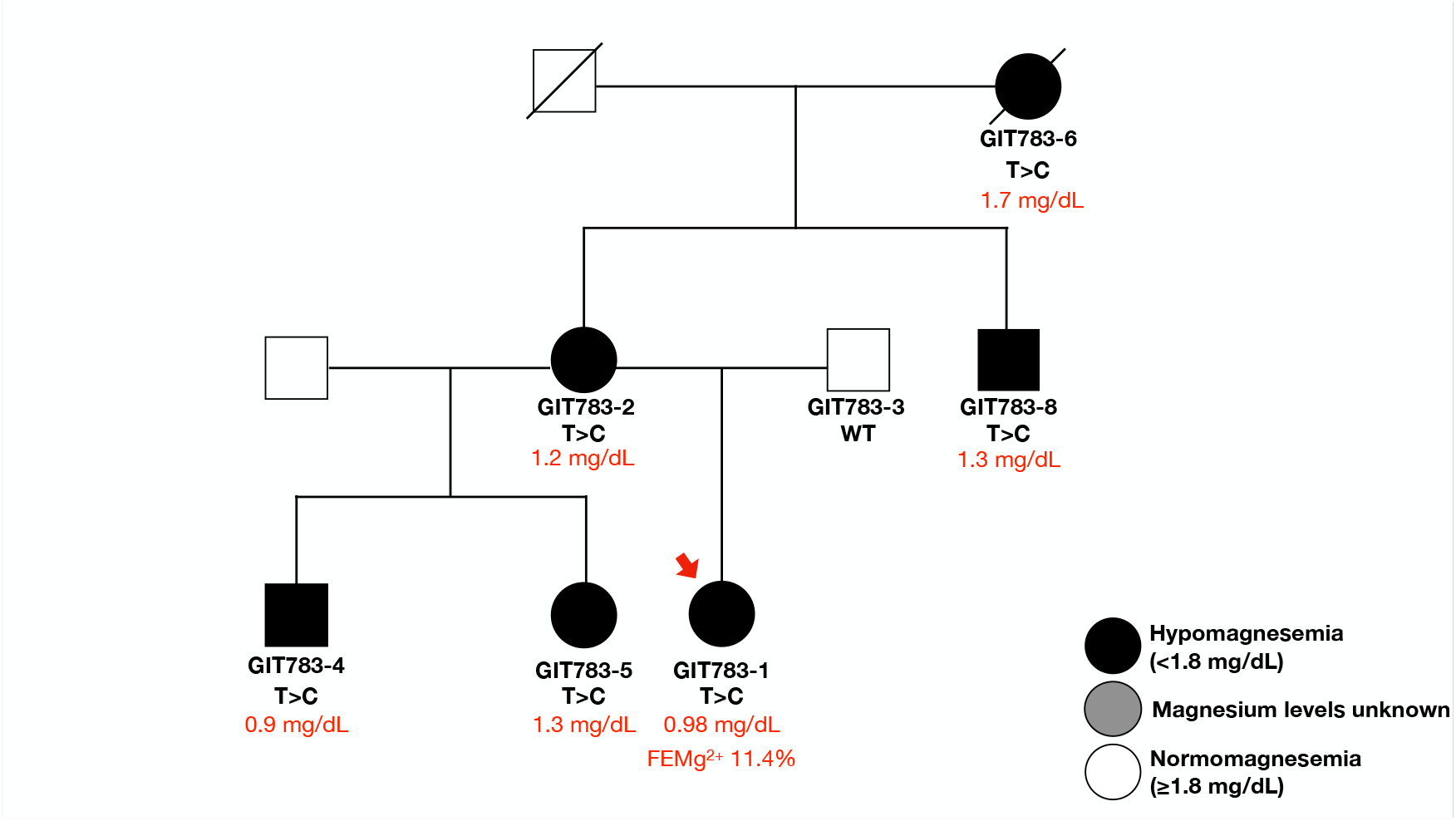
Kindred GIT783 family with hypomagnesemia. Partial pedigree of kindred GIT783. Proband GIT783-1 is shown with a red arrow. Serum magnesium and m.4290 genotypes are shown below each kindred member.

A defect in mitochondrial respiratory chain activity is a leading hypothesis for how these mutations result in a Gitelman-like phenotype. Several groups have reported a reduction in activity in mitochondrially-encoded respiratory complex subunits as a hallmark of pathological mitochondrial tRNA mutations^39,40,41,42^. One study found mitochondria with distorted morphology in the distal tubule in kidney biopsies from patients with homoplasmic MT-tRNA mutations, which was not seen in mitochondria from the proximal tubule and other tissues^41^. Importantly, the anatomic distal tubule includes both the collecting duct and distal convoluted tubule, the latter of which is involved in renal magnesium reabsorption. Altogether, these findings shed light on how mitochondrial pathology can contribute to renal disease and salt-wasting phenotypes.

Interestingly, this m.4290 U>C homoplasmic mutation has been previously reported once in three daughters, who were described as having progressive necrotizing encephalopathy^39^. Unfortunately, no electrolyte chemistries were reported in this study. Notably, all but one carrier of m.4290 U>C in kindred GIT783 suffers from major depressive disorder; the exception is GIT783-1’s half sibling who suffers from anxiety. Further research will be needed to understand how/whether this variant is contributing to neurologic phenotypes and whether this is directly related to hypomagnesemia or other mechanisms related to mitochondrial function.

### 17q12 deletion in patients with hypomagnesemia

Among six other patients with unexplained hypomagnesemia, a structural variant analysis revealed that *HNF1B* was completely deleted in each case (**Fig. S15A**). The evidence included loss of heterozygosity for all SNPs with MAF 0.2 to 1.0, spanning a 1.26 Mb segment of chromosome 17q12 that includes HNF1B, consistent with a monoallelic deletion. This pattern was not seen in any of the other 381 subjects (**Fig. S15B**). Heterozygous LOF mutations in HNF1B cause hypomagnesemia and maturity onset diabetes of the young (MODY)^43,44,45^.

Deletion of 17q12 is associated with myriad phenotypes, including MODY, structural kidney abnormalities, hypomagnesemia, neuropsychiatric and neurodevelopmental disorders, with renal patholgy being the most prevalent finding at an incidence of 90% in cases. Interestingly, only one proband in the present cohort suffered from diabetes and was prescribed metformin for glycemic control.^46^ Interestingly, half of these patients were diagnosed with major depressive disorder and were reported as taking antidepressants.

### Biallelic mutations in other salt wasting genes

Among the remaining patients with clinical criteria for salt-wasting disorders hallmarked by hypokalemia and/or hypomagnesemia, biallelic damaging mutations of the five Bartter’s syndrome genes, including 36 in *CLCNKB*, 14 in *SLC12A1*, 7 in *KCNJ1*, 3 in *BSND*, and 2 in *MAGED2* mutations,as well as 2 in CLDN10, 1 in KCNJ10, and 2 in SLC26A3 (linked to congenital chloride diarrhea)^22^. Here too, there was strong consistency of phenotypic features with those expected to result from a mutation in each gene. Overall, patients with mutations in Bartter syndrome genes presented in the first years of life, had high rather than low urinary calcium, and had higher Mg^2+^ levels than found in GS. (**Fig. 2**)

Importantly, expanding our analysis to include rare intronic mutations detected by WES in BS patients identified one noncoding mutation in a noncanonical splice site in SLC12A1. This patient, GIT703-1, harbors a frameshift mutation in SLC12A1; a second mutation lies 6 basepairs upstream of the intron-exon boundary of exon 5 (**Fig. S16A**,**B**). SpliceAI predicts with high confidence (score = 0.98) that this mutation creates a novel splice acceptor site, likely due to the enriched polypyrimidine tract immediately upstream, which favors the new site over the wild-type location (**Fig. S16C**). A complete list of all biallelic variants found in all these probands can be found in **Table S7**.

## Discussion

Analysis of 329 probands referred for suspected renal electrolyte disorders identified likely pathogenic genotypes in 83% of probands. The conclusion that identified genotypes are pathogenic is supported by the electrolyte values found in patients with mutations in each gene. These genotypes included 200 biallelic mutations in *SLC12A3* found by WES or WGS-the latter included two deep intronic splice gains and an exonic deletion missed by WES. Virtually all patients with the combination of hypokaemia, hypomagnesemia and hypocalciuria had biallelic *SLC12A3* mutations. Among 60 patients with biallelic mutations in Bartter syndrome genes, normal or hypercalciuria with normal magnesium levels predominated. Likely disease-causing mutations in *HNF1B, MAGED2, CLDN10* and mitochondrial DNA were much less common and also associated with characteristic electrolyte values.

Among patients with zero mutations in known genes identified, none of fifteen with K+ values had hypokalemia, 5 had isolated hypomagnesemia and 14 others were referred without lab values. Twenty-three other patients had monoallelic variants in genes other than *SLC12A3*, leaving the question of whether other recessive alleles might be identified by WGS in these patients. No evidence for digenic mutations (trans compound heterozygotes) was identified.

The identification of likely disease-related mutations that lie outside canonical GT/AG splice donor and acceptor sites that are captured by WES highlight the importance of analyzing the full span of sequences that influence splicing efficiency. Such mutations accounted for less than 10% of splice-altering mutations, less than what has been reported for Fabry disease.^47^ In addition, we found that GATK gCNV provides a robust tool capable of capturing nearly 90% of small deletions in genes linked to Mendelian renal salt-wasting disorders.

To date, only a single deep intronic mutation in SLC12A3 that introduces a new splice donor site and a resulting cryptic exon (found twice) was found in Gitelman syndrome. Our findings here suggest that these non-coding pathogenic alleles are scarce but can be critical for clinical diagnosis in select patients.

Thus collectively, among 400 mutant SLC12A3 alleles in patients with biallelic mutations, only two instances of a heterozygous deep intronic splice gain were identified. Five other disease-related alleles are unaccounted for but have not been identified by WES or WGS. Thus for *SLC12A3*, the fraction of mutations lying in non-coding regious beyond the range of WES is between 0.5% and 2%.

This study also identified a proband with severe hypomagnesemia and a novel homoplasmic mutation the mitochondrial gene encoding its isoleucine tRNA. The mutated base lies two bases from the tRNA’s anticodon loop and is at a position completely conserved from yeast to humans. Study of other family members of the newly ascertained family showed that hypomagesemia is inherited on the maternal lineage, along with the mitochondrial mutation, in this family. Interestingly, mutation carriers in this family had signifcant histories of depression or anxiety. We previously described a very large family with maternally inherited hypomagnesemia with a T > C mutation at the adjacent base that also resulted in renal hypomagnesemia and was passed exclusively from mothers to their offspring. Similar homoplasmic mutations in the mitochondrial tRNA^ile^ associated with hypomagnesemia have recently been reported.^40^

## Data Availability

All data produced in the present study are available upon reasonable request to the authors

