## Supplemental Figures and Tables for "Mutations in 329 probands with suspected renal electrolyte disorders"

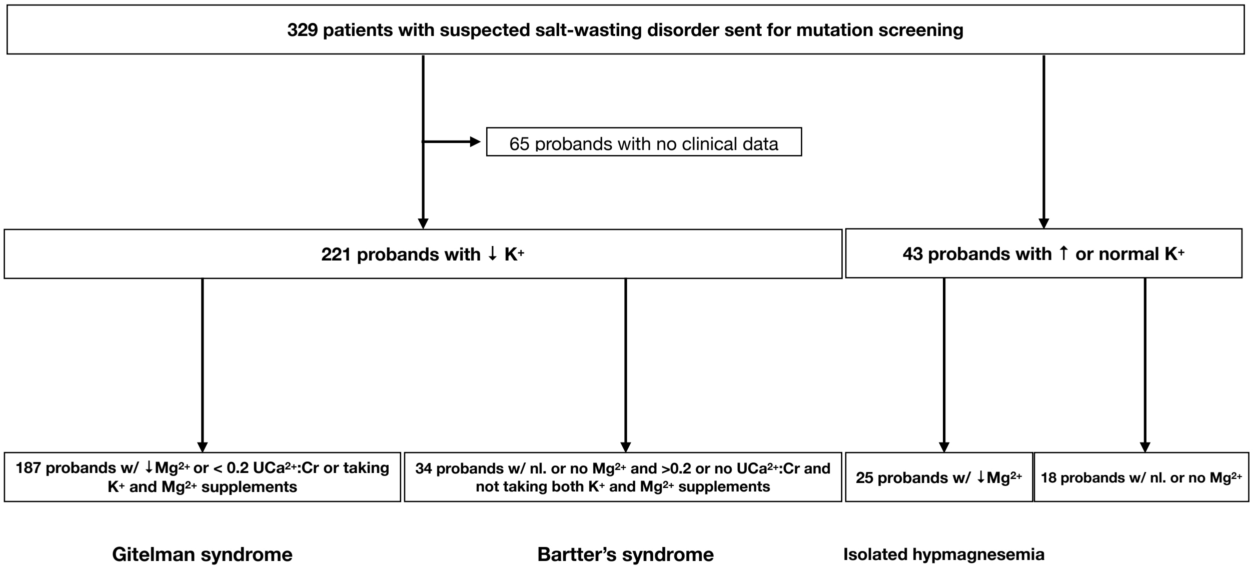


**Fig. S1. Overview of phenotypes in salt-wasting cohort.** Phenotypes were split based off different serum electrolyte cutoffs. A cutoff of <3.5 mmol/L for $K^+$ and <1.8 mg/dL for $Mg^{2+}$ were used. $UCa^{2+}$:Cr indicates urinary calcium to creatinine ratio.

**A**

| PID | chr | pos | ref | alt | gene | mut | consequence | gnomAD WGS | CADD20 | BP | $K^+$ | $Mg^{2+}$ | $Ca^{2+}$:Cr |
|---|---|---|---|---|---|---|---|---|---|---|---|---|---|
| GIT418-1 | 16 | 56947166 | T | A | SLC12A3 | c.2952-10T>A | splice loss | 0 | 22.5 | 120/80 | 3.2 | 0.64 | - |

**B**

| variant | gene | Δ type | Δ score | pre-mRNA position |
|---|---|---|---|---|
| 16-56947166-T-A<br>UCSC, gnomAD | SLC12A3 (ENSG00000070915.10_1 / ENST00000566786.5_1 / NM_001126107.2)<br>biotype: protein coding<br>OMIM, GTEx, gnomAD, ClinGen, Ensembl, Decipher, GeneCards | Acceptor Loss | 0.82 | 10 bp |
| | | Donor Loss | 0.00 | |
| | | Acceptor Gain | 0.72 | 2 bp |
| | | Donor Gain | 0.00 | |

**C**

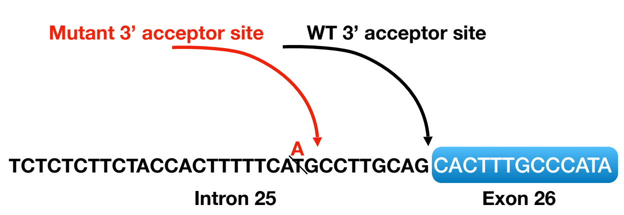


**D**

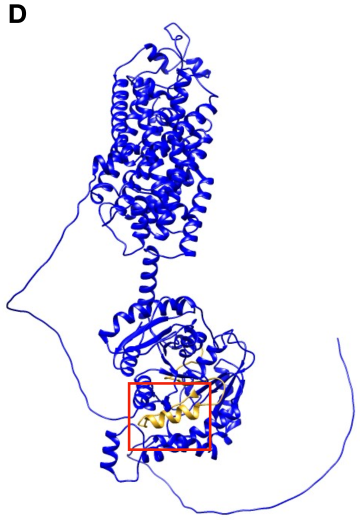

**Fig. S2. Intronic mutation upstream of splice acceptor site introduces frameshift in exon 26.** (A) Information on variant c.2952-10T>A that results in a frameshift mutation at the 5' end of exon 26 followed by clinical features of GS proband (B) SpliceAI webtool identifies variant c.2952-10T>A as a disruptor of exon 26's splice acceptor site (C) Illustration of the noncoding variant in polypyrimidine tract upstream of splice accepter site and resulting truncated protein product caused by a frameshift in 5’ end of exon 26 (D) Highlighed in red is the location of the deleted residues that result from the frameshift introduced by this variant.

**A**

| PID | chr | pos | ref | alt | gene | mut | consequence | gnomAD WGS | CADD20 | BP | K+ | Mg2+ | Ca2+:Cr |
|---|---|---|---|---|---|---|---|---|---|---|---|---|---|
| GIT398-1 | 16 | 56938379 | G | A | SLC12A3 | c.2948+5 AG>AA | +5 Ex-In boundary | 0 | 24.3 | 120/75 | 2.8 | 1.22 | 0.09 |

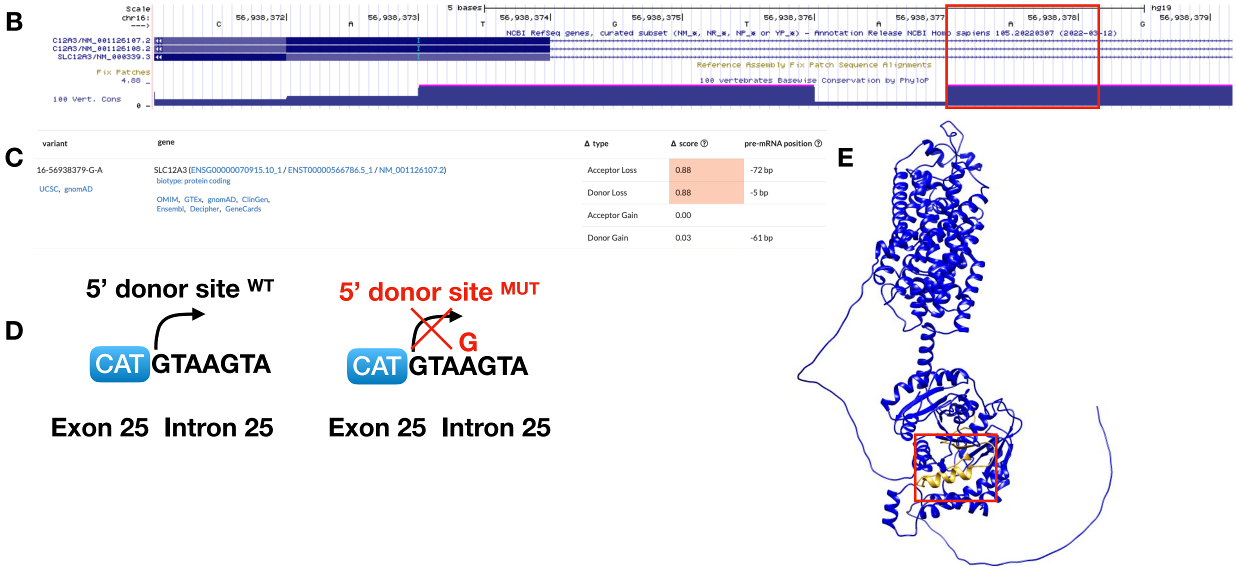


**Fig. S3. Non-coding variant downstream of 3’ end of exon 25 in SLC12A3 results in loss of splice donor site.** (A) Clinical features of GS proband and information on noncoding variant in nearly invariant donor site downstream of dinucleotide motif. (B) View of noncoding variant on UCSC genome. Conservation scores located at the bottom show that this postion is nearly invariant. (C) SpliceAI predicts the loss of this native donor site at this location with high confidence (score = 0.88, high precision cutoff is >0.8). (D) Illustration by how this noncoding variant in a noncanonical splice site disrupts splicing. (E) Loss of donor site effectively removes last 46 amino acids in SLC12A3 C-terminus (yellow). Structure generated from Alphafold prediction software.

**A**

| PID | chr | pos | ref | alt | gene | mut | consequence | gnomAD WGS | CADD20 | BP | $K^{+}$ | $Mg^{2+}$ | $Ca^{2+}$:Cr |
|---|---|---|---|---|---|---|---|---|---|---|---|---|---|
| GIT699-1 | 16 | 56920391 | A | G | SLC12A3 | c.2034+4A>G | +4 Ex-In boundary | 3.99E-06 | 23.7 | 116/63 | 2.1 | 1.8 | 0.007 |

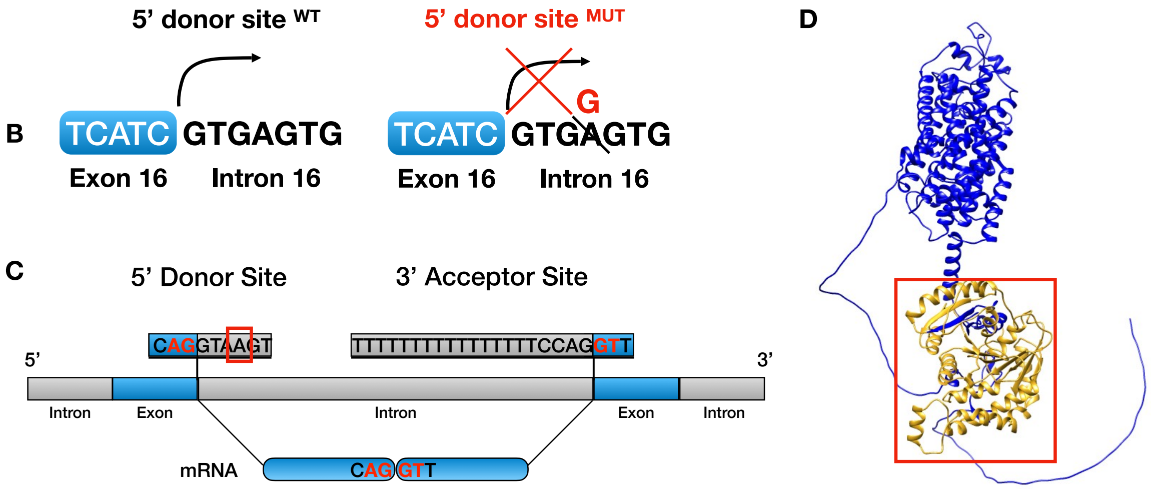


**Fig. S4. Non-coding variant downstream of 3' end of SLC12A3 exon 16 results in loss of splice donor site.** (A) Clinical features of GS proband and information on noncoding variant in nearly invariant donor site downstream of dinucleotide motif. (B) Illustration demonstrating how noncoding variant 16-56920391 A>G results in disruption of the extended 5' splice donor site sequence 'GTAAGT' at Exon 16. (C) Sequence specificity at the invariant dinucleotide motif (red) and noncanonical splice sequences (black) at the exon-intron boundary. Location of 16-56920391 A>G is highlighted in a red box. (D) Loss of donor site effectively removes c-terminal half of SLC12A3 (yellow). Structure generated from Alphafold prediction software.

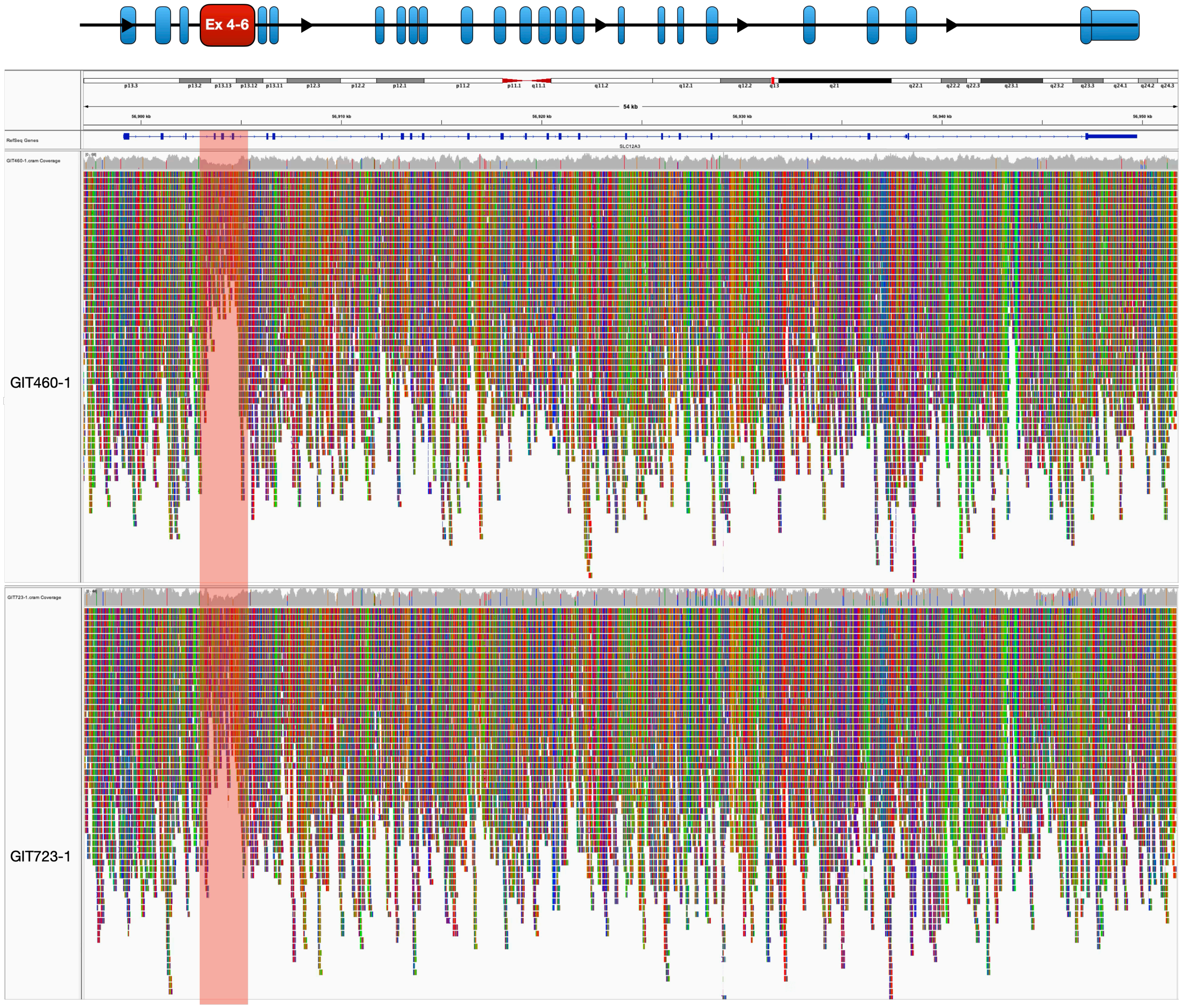


**Fig. S5. Visualization of SLC12A3 intragenic deletions in probands GIT460-1 and GIT723-1 using IGV.** (A) Aligned sequencing reads mapping to SLC12A3 for probands GIT460-1. Aligned sequencing reads mapping to SLC12A3 for probands GIT460-1 and GIT723-1. Deletions shown in red. A heterozygous deletion can be seen based on the reduced coverage within the region in red relative to all other locations on the gene.

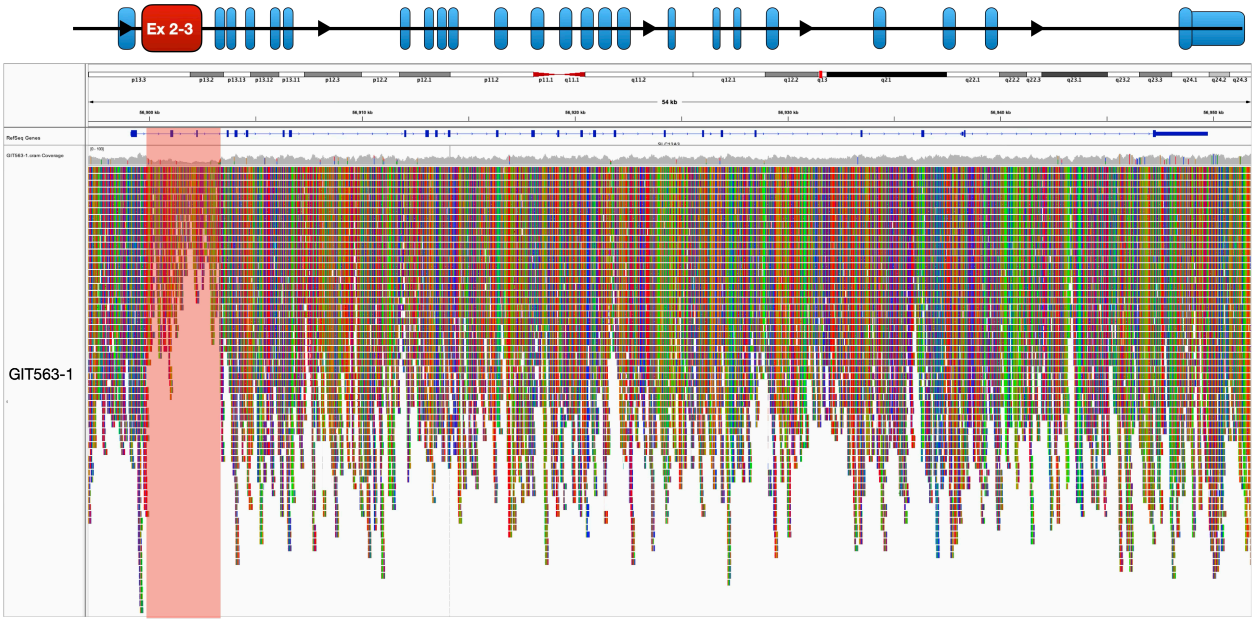


**Fig. S6. Visualization of SLC12A3 intragenic deletions in proband GIT563-1 using IGV.** (A) Aligned sequencing reads mapping to SLC12A3 for probands GIT563-1. Deletions are shown in red. A heterozygous deletion can be seen based on the reduced coverage within the region in red relative to all other locations on the gene.

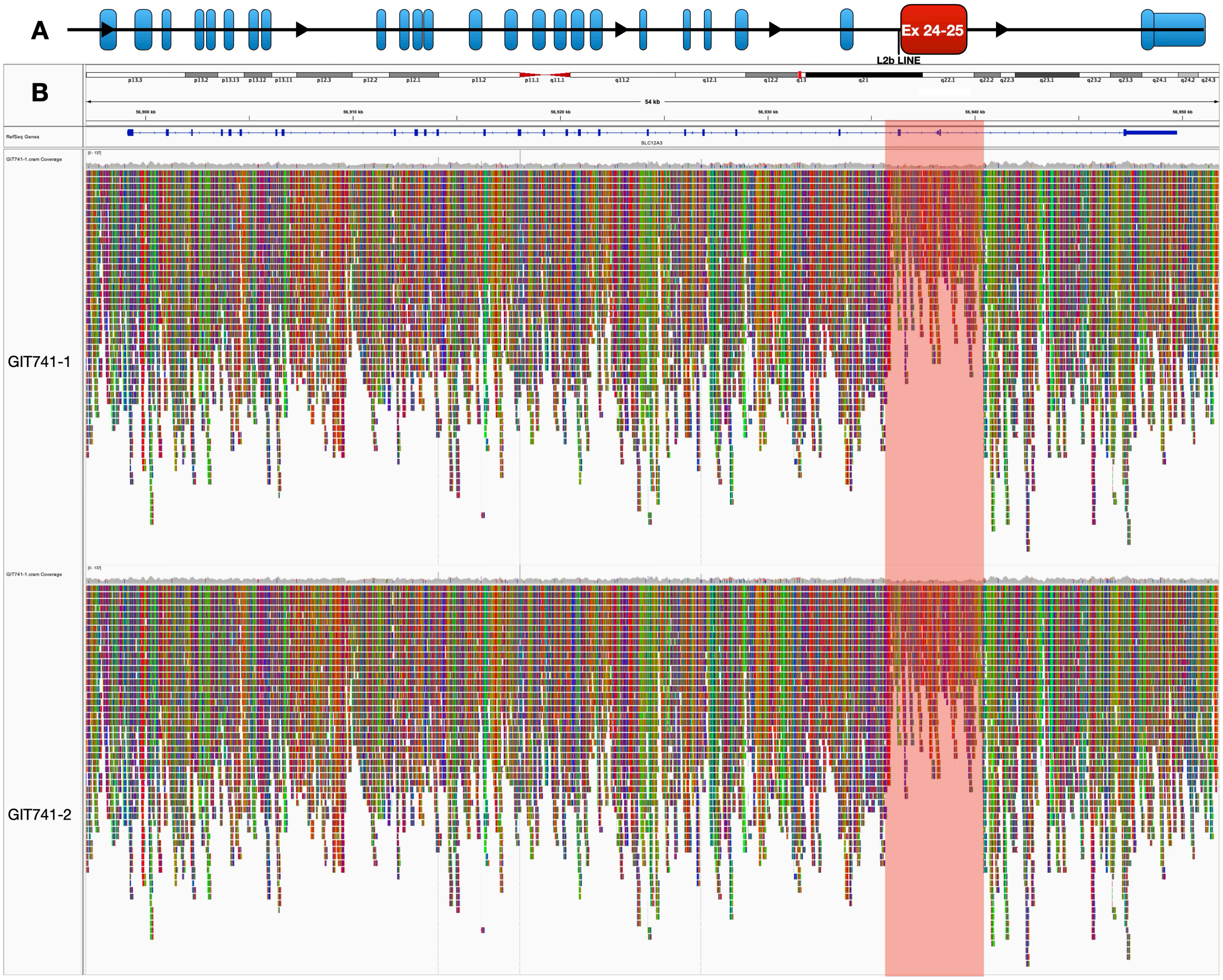


**Fig. S7. Visualization of SLC12A3 intragenic deletion of exons 24 and 25 in probands GIT741-1 and GIT742-2 using IGV.** (A) Illustration of intragenic deletion of exons 24 and 25 in GS fraternal twin probands GIT741-1 and GIT741-2. (B) IGV view of aligned WGS sequencing reads mapped to SLC12A3 in GIT741-1 and 741-2. A heterozygous intragenic deletion of SLC12A3 can be seen by the reduced sequencing coverage at exons 24 and 25 (red) relative to all other regions in the gene.

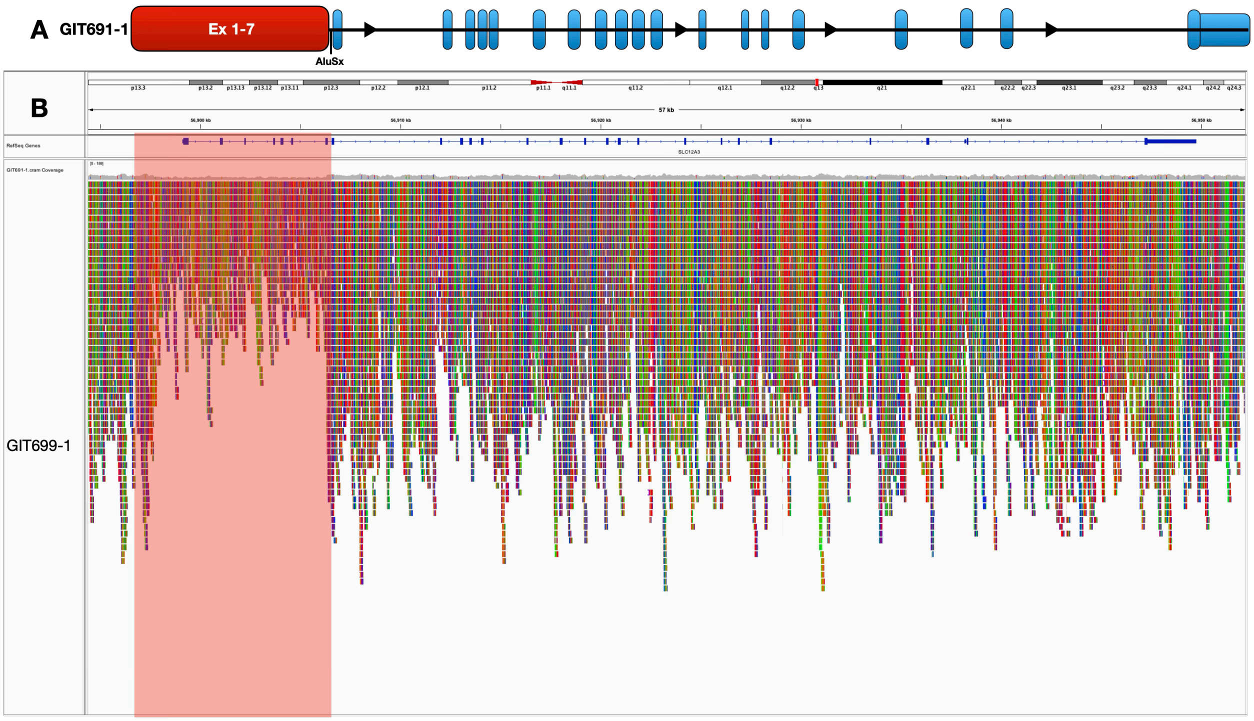


**Fig. S8. Visualization of SLC12A3 intragenic deletion of exons 1-7 in proband GIT691-1 using IGV.** (A) Illustration of intragenic deletion of exons 1-7 in GS proband GIT691-1. (B) IGV view of aligned WGS sequencing reads mapped to SLC12A3 in GIT691-1. A heterozygous intragenic deletion of SLC12A3 can be seen by the reduced sequencing coverage between ~2 kb upstream of SLC12A3 and intron 7 (red) relative to all other regions in the gene.

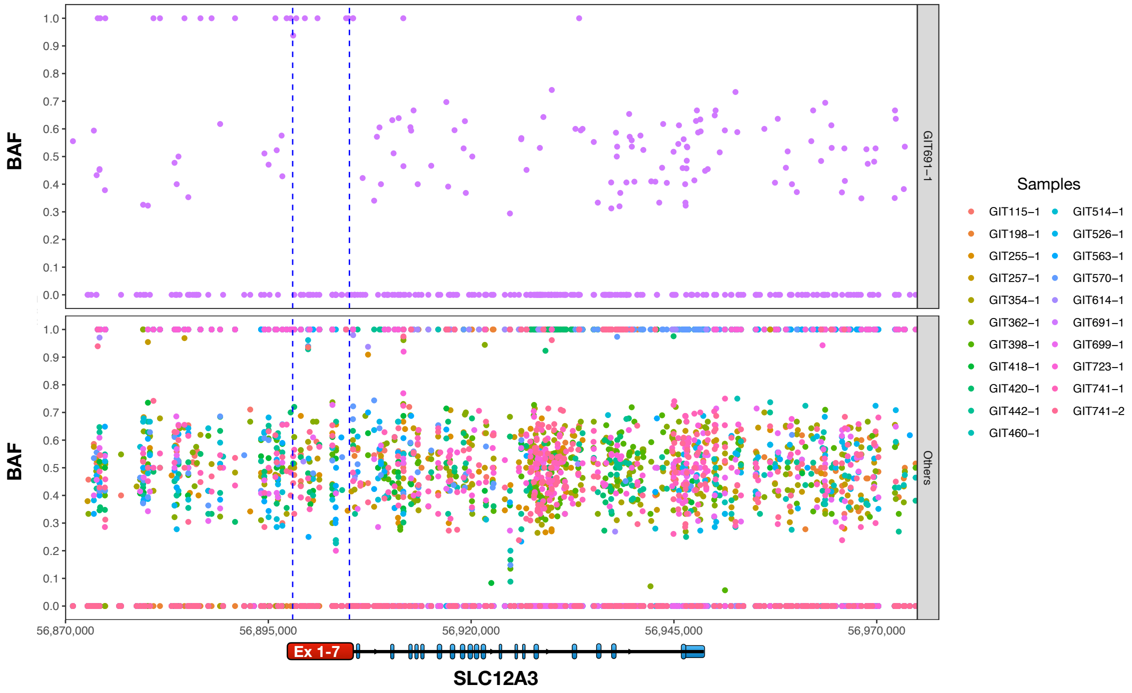


**Fig. S9. B-allele frequency analysis of SLC12A3 in proband GIT691-1.** The allele frequencies of common variants (MAF >0.2 and <0.8) within chr16:56,870,000-56,975,000 (hg19) are plotted on the y-axis and genomic location on the x-axis. A 9 kb region containing no common heterozygous variants is seen in GIT691-1 at the site of the putative deletion. A schematic of the SLC12A3 is shown at the bottom for reference.

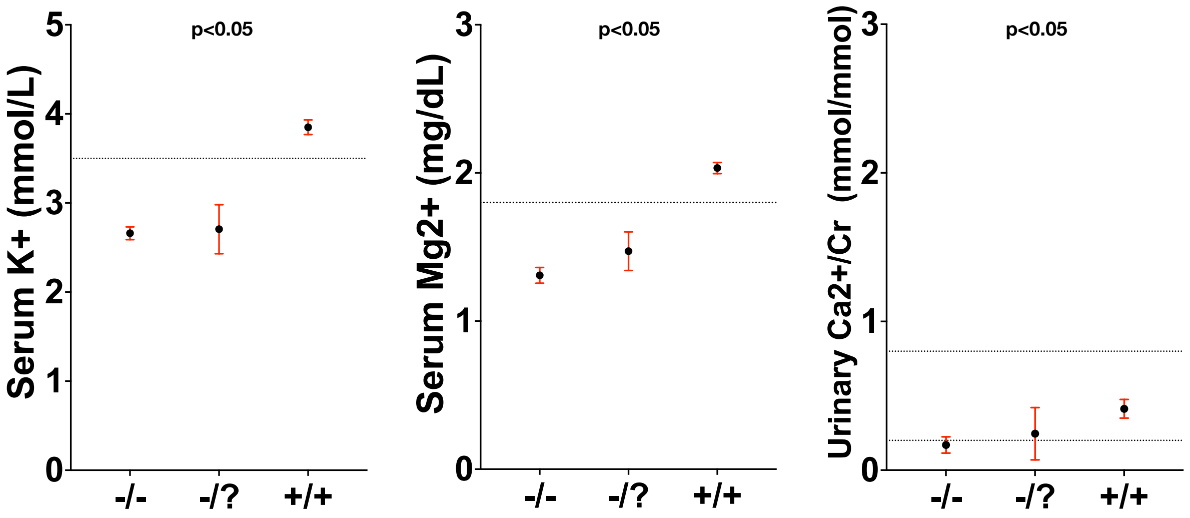

**Fig. S10. SLC12A3 probands with deleterious monoallelic mutations phenocopy SLC12A3 homozygotes.** Effect of SLC12A3 mutations on serum laboratory parameters. Each black dot represents the mean. The mean ± SEM values of indicated laboratory parameters are shown for individuals inheriting 0 (+/+), 1 (+/?), or 2 (−/−) coding mutations in SLC12A3. Values for serum Mg2+ can be converted to mmol/L by multiplying by 0.4114. Cr indicates creatinine. P values among different genotype classes are shown. The size of each group is annotated at the bottom.

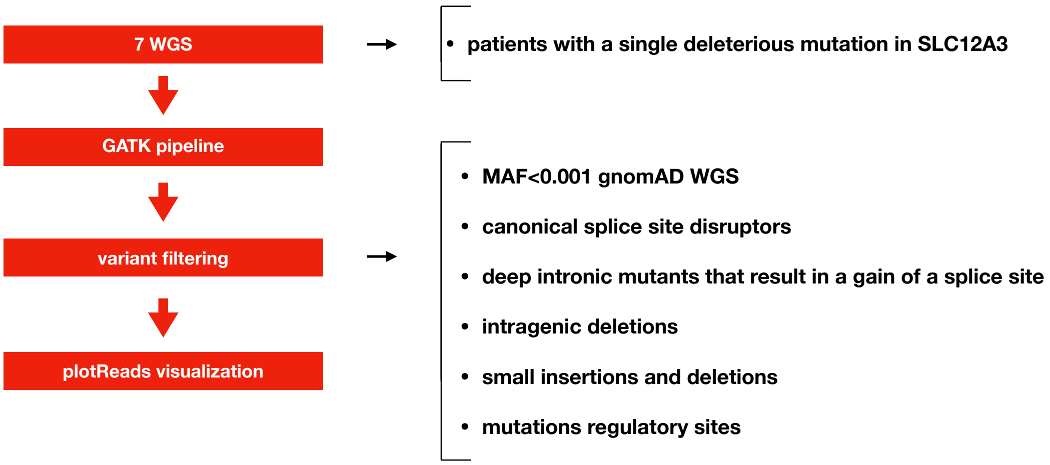


**Fig. S11. Lifton lab bioinformatic pipeline for noncoding variant identification.** Outline of steps used to identify deleterious mutations within the noncoding genome of 7 salt-wasting

**A**

| PID | chr | pos | ref | alt | gene | mut | consequence | gnomAD WGS | CADD20 | BP | $K^+$ | $Mg^{2+}$ | $Ca^{2+}$:Cr |
|---|---|---|---|---|---|---|---|---|---|---|---|---|---|
| GIT250-1 | 16 | 56917770 | C | T | SLC12A3 | c.1670-191GC>GT | splice gain | 3.19E-05 | 27 | 120/78 | 3 | 1.5 | 0.08 |
| GIT255-1 | 16 | 56917770 | C | T | SLC12A3 | c.1670-191GC>GT | splice gain | 3.19E-05 | 27 | 114/74 | 2.6 | 0.87 | 0.07 |

**B**

| Variant | Gene | Δ type | Δ score | position |
|---|---|---|---|---|
| 16-56917770-C-T | SLC12A3 ( ENSG00000070915.10_16 / ENST00000566786.5_7 / NM_001126107.2) | Acceptor Loss | 0.00 | |
| intron variant | protein coding (plus strand) | Donor Loss | 0.05 | 44 bp |
| UCSC, gnomAD | OMIM, GTEx, gnomAD, ClinGen, Ensembl, Decipher, GeneCards | Acceptor Gain | 0.13 | -37 bp |
| | | Donor Gain | 0.72 | -2 bp |

**Fig. S12. Two patients with identical deep intronic mutations known to introduce a cryptic exon between exons 13 and 14.** (A) Information on c.1670-191GC>GT variant in intron 13 of SLC12A3 followed by clinical features of two GS probands. (B) SpliceAI webtool identifies variant c.1670-191GC>GT as the gain of a splice donor site that has been show to introduce a cryptic exon between exons 13 and 14(score = 0.72, recommended cutoff is >0.5).

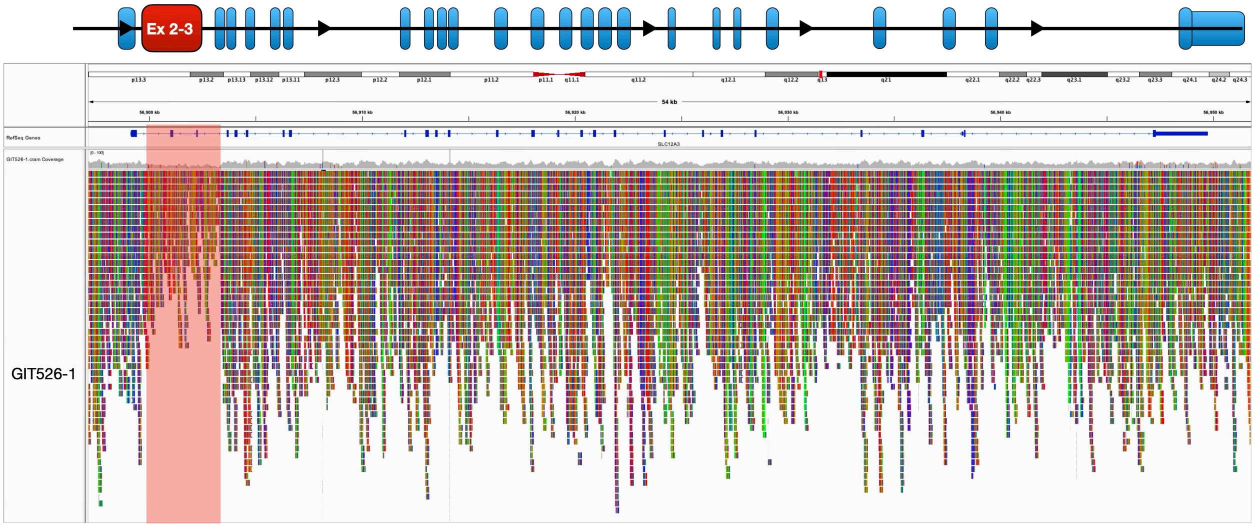


**Fig. S13. Visualization of SLC12A3 intragenic deletions in probands GIT526-1 using IGV.** (A) Aligned sequencing reads mapping to SLC12A3 for probands GIT526-1. Deletions are shown in red. A heterozygous deletion can be seen based on the reduced coverage within the region in red relative to all other locations on the gene.

**A**

| PID | chr | pos | ref | alt | gene | mut | consequence | gnomAD WGS | CADD20 | BP | K+ | $Mg^{2+}$ | $Ca^{2+}$:Cr |
|---|---|---|---|---|---|---|---|---|---|---|---|---|---|
| GIT336-1 | 16 | 56900664 | G | A | SLC12A3 | c.283-318G>A | regulatory mutation | 0 | 3.4 | 140/90 | 2.6 | 1.24 | 0.02 |

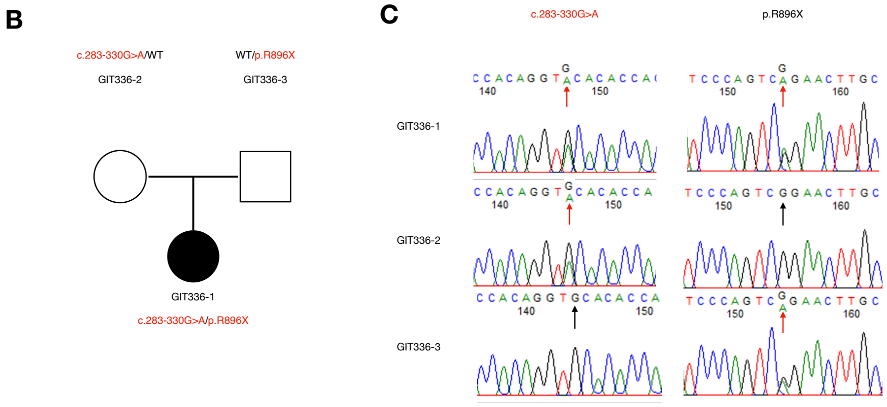


**D**

| Species | Base |
|---|---|
| Homo Sapien | G |
| Chimp | G |
| Gorilla | G |
| Orangutan | G |
| Gibbon | G |
| Rhesus | G |
| Macaque | G |
| Baboon | G |
| Monkey | G |
| Marmoset | G |
| Bushbaby | G |
| Squirrel Monkey | G |

**Fig. S14. Variant of unknown function located in SLC12A3 intron 1.** (A) Phenotypic features of GIT336-1 and information on rare noncoding variant within intron 1 of SLC12A3. (B) Pedigree for Gitelman proband GIT336-1 showing noncoding mutation c.283-318G>A was inherited from one parent (GIT336-2) and pathogenic coding mutation p.R896X was inherited from the other parent (GIT336-3). (C) Targeted sequencing showing SLC12A3 noncoding mutation c.283-318G>A and pathogenic coding mutation p.R896X. Black arrows point to wild-type alleles and red arrows point to mutant alleles. (D) Table demonstrating 100% conservation at loci 16-56900664 within primates.

**A**

| PID | count | chr | start | end | consequence | gene | gnomAD SV | BP | K+ | Mg2+ | UCa2+:Cr |
|---|---|---|---|---|---|---|---|---|---|---|---|
| GIT484-1 | 6 | 17 | 34842294 | 36105125 | 1.26 Mb del | HNF1B | 1.40E-04 | 110/70 | 2.60 | 0.50 | 1.3 |
| GIT487-1 | 6 | 17 | 34842294 | 36105125 | 1.26 Mb del | HNF1B | 1.40E-04 | 117/58 | 3.00 | 0.80 | - |
| GIT499-1 | 6 | 17 | 34842294 | 36105125 | 1.26 Mb del | HNF1B | 1.40E-04 | - | 3.85 | 1.39 | - |
| GIT670-1 | 6 | 17 | 34842294 | 36105125 | 1.26 Mb del | HNF1B | 1.40E-04 | 110/98 | 3.40 | 1.00 | - |
| GIT696-1 | 6 | 17 | 34842294 | 36105125 | 1.26 Mb del | HNF1B | 1.40E-04 | - | 3.50 | 1.10 | - |
| GIT728-1 | 6 | 17 | 34842294 | 36105125 | 1.26 Mb del | HNF1B | 1.40E-04 | 110/70 | 3.50 | 1.20 | - |

**B**

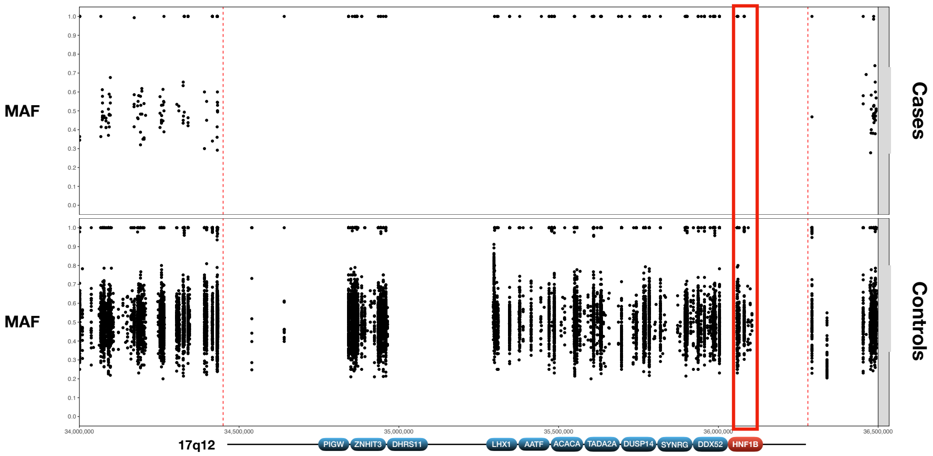


**Fig. S15. 17q12 deletion in 6 salt-wasting probands with hypomagnesemia.** (A) Clinical information and mutation of the 17q12 monoallelic deletion in six patients with hypomagnesemia is shown. (B)The allele frequencies of common variants (MAF >0.2 and <0.8) within 17q12 are plotted on the y-axis and genomic location on the x-axis. In the top panel, there is a loss of heterozygosity for all six patients across a 1.26 Mb region that includes HNF1B, consistent with the deletion of this segment. In contrast, in the bottom panel MAF of SNPs in genes in 323 WT subjects are shown, showing a mean close to 50%, consistent with heterozygosity of these variants.

**A**

| PID | chr | pos | ref | alt | gene | mut | consequence | gnomAD WGS | CADD20 | BP | K+ | Mg2+ | Ca2+:Cr |
|---|---|---|---|---|---|---|---|---|---|---|---|---|---|
| GIT703-1 | 15 | 48518043 | A | G | SLC12A1 | c.629-6A>G | splice loss | 6.2E-06 | 16.2 | 63/43 | 4.6 | 1.5 | - |

**B**

| Variant | Gene (= MANE Select transcript; = non-coding transcript) | Δ type | Δ score | position | REF score | ALT score |
|---|---|---|---|---|---|---|
| 15-48518043-A-G | SLC12A1 ( ENSG00000074803.21_17 / ENST00000380993.8_9 / NM_000338.3) | Acceptor Loss | 0.22 | 6 bp | 0.22 | 0.00 |
| non coding transcript exon variant | protein coding MANE Select transcript (plus strand) | Donor Loss | 0.00 | | 0.00 | 0.00 |
| UCSC, gnomAD | OMIM, GTEx, gnomAD, ClinGen, Ensembl, Decipher, GeneCards | Acceptor Gain | 0.98 | 1 bp | 0.00 | 0.98 |
| | | Donor Gain | 0.00 | | 0.00 | 0.00 |

**C**

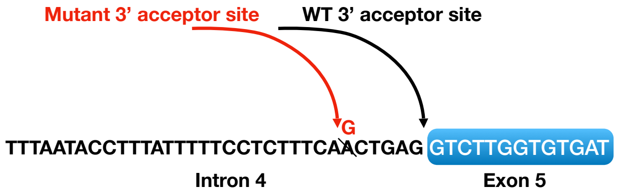


**Fig. S16. Intronic mutation upstream of splice acceptor site introduces frameshift in exon 5 of SLC12A1.** (A) Information on variant c.629-6A>G that results in a frameshift mutation at the 5' end of exon 5 followed by clinical features of BS proband (B) SpliceAI webtool identifies variant c.2952-10T>A as a disruptor of exon 5’s splice acceptor site (C) Illustration of the noncoding variant in polypyrimidine tract upstream of splice accepter site and resulting truncated protein product caused by a frameshift in 5’ end of exon 5

| PID | BP | K+ | Mg2+ | HCO3- | UCa2+/Cr | Gender | Medications |
|---|---|---|---|---|---|---|---|
| GIT110-1 | 110/60 | 3.00 | 1.22 | 31.00 | 0.08 | F | - |
| GIT123-1 | 120/60 | 2.60 | 1.27 | 32.00 | 0.01 | M | KCl |
| GIT125-1 | 120/80 | 2.40 | 1.71 | 27.00 | 0.03 | M | amiloride |
| GIT127-1 | - | - | - | - | - | F | - |
| GIT128-1 | 100/64 | 2.80 | 2.20 | 29.00 | 0.00 | M | Kcl |
| GIT129-1 | 110/70 | 2.60 | 1.30 | 30.50 | 0.47 | M | - |
| GIT147-1 | 80/60 | 2.60 | 1.10 | 36.00 | 0.83 | F | KCl |
| GIT150-1 | 80/50 | 2.10 | 1.70 | 30.00 | 0.34 | F | - |
| GIT151-1 | 80/60 | 2.80 | 1.68 | - | 0.31 | F | Kcl, spirinolactone, |
| GIT169-1 | - | - | - | - | - | F | - |
| GIT171-1 | - | - | - | - | - | F | - |
| GIT172-1 | 100/60 | 2.80 | 0.32 | 29.00 | 0.22 | F | KCl, Mg |
| GIT176-1 | - | - | - | - | - | F | - |
| GIT180-1 | - | - | - | - | - | F | - |
| GIT182-1 | 110/70 | 1.90 | 1.87 | 24.00 | 1.10 | M | - |
| GIT191-1 | 96/60 | 3.50 | 1.87 | 28.00 | - | F | indocin |
| GIT192-1 | - | - | - | - | - | M | - |
| GIT193-1 | 120/80 | 2.80 | 0.80 | 30.00 | 0.00 | M | Mg, indocin |
| GIT201-1 | - | 2.10 | 1.70 | 37.40 | - | F | spironolactone |
| GIT214-1 | 110/60 | 2.30 | 1.79 | 30.00 | - | M | K, spironolactone, aspirin |
| GIT217-1 | - | - | - | - | - | F | - |
| GIT235-1 | - | 3.50 | 1.30 | 24.00 | - | F | - |
| GIT240-1 | - | - | - | - | - | F | - |
| GIT250-1 | 88/54 | 2.20 | 1.50 | 28.00 | 0.08 | M | Kcl, amiloride |
| GIT255-1 | 114/74 | 2.60 | 0.87 | 31.00 | 0.07 | M | Kcl, amiloride, Mg |
| GIT257-1 | 110/59 | 2.80 | 2.31 | 32.00 | 1.20 | M | Kcl, amiloride, growth |
| GIT259-1 | - | - | - | - | - | M | - |
| GIT266-1 | 90/60 | 3.80 | 2.10 | 26.50 | 0.28 | F | K citrate |
| GIT275-1 | 138/76 | 3.50 | 1.99 | 29.00 | 0.06 | M | Kcl |
| GIT278-1 | - | - | - | - | - | M | KCl |
| GIT280-1 | - | 2.90 | 1.44 | - | 0.00 | M | KCl |
| GIT287-1 | 77/38 | 2.70 | 1.30 | 29.00 | 1.92 | F | Kcl, spironolactone, |
| GIT293-1 | 118/74 | 3.00 | 1.90 | 33.00 | 0.07 | M | KCl, indocin, amiloride |
| GIT295-1 | - | - | - | - | - | F | - |
| GIT296-1 | 91/45 | 3.30 | 1.10 | 24.70 | 2.80 | F | Kcl, indocin |
| GIT299-1 | - | 4.00 | 1.82 | 23.50 | - | F | indocin |
| GIT301-1 | - | 2.70 | 1.17 | - | 0.05 | F | - |
| GIT311-1 | 136/68 | 3.60 | 1.70 | 32.00 | 0.14 | M | KCl, amiloride |
| GIT318-1 | 92/43 | 3.90 | 2.04 | 26.00 | 4.59 | M | Kcl, indocin |
| GIT325-1 | 90/70 | 2.00 | 1.81 | 28.00 | - | F | K |
| GIT326-1 | 110/75 | 2.80 | 2.00 | 38.00 | 0.04 | M | indocin |
| GIT328-1 | 110/80 | 2.70 | 1.20 | 27.00 | 0.03 | F | Mg |
| GIT331-1 | - | - | - | - | - | F | - |
| GIT332-1 | 99/66 | 2.30 | 2.82 | 26.00 | 4.20 | F | indocin |
| GIT336-1 | 140/90 | 2.60 | 1.20 | 32.00 | 0.02 | M | Kcl, Mg |
| GIT343-1 | 120/78 | 3.50 | 1.22 | - | - | F | - |
| GIT348-1 | 110/60 | 2.90 | 1.41 | 31.00 | 0.06 | F | Kcl, Mg, indocin |
| GIT360-1 | 110/80 | 3.30 | 1.50 | 32.00 | 0.60 | F | Kcl, indocin, Mg |
| GIT369-1 | 120/90 | 2.50 | 1.14 | 29.00 | 0.01 | F | Mg |
| GIT374-1 | - | 3.20 | - | 28.00 | 2.33 | F | - |
| GIT377-1 | 128/78 | 3.20 | 0.90 | 31.00 | 0.05 | F | Kcl, Mg, Amiloride |
| GIT379-1 | 120/70 | 3.10 | 1.14 | - | 0.15 | F | Kcl, Mg |
| GIT384-1 | 120/78 | 2.80 | 1.70 | 27.00 | 0.06 | F | Kcl |
| GIT387-1 | 143/73 | 2.40 | 1.00 | 31.00 | 0.00 | F | - |
| GIT390-1 | 140/80 | 4.60 | - | 27.00 | 0.80 | M | Kphos |
| GIT392-1 | 110/70 | 3.00 | 1.22 | 27.00 | - | F | K |
| GIT393-1 | 135/85 | 3.60 | 1.40 | 20.00 | - | F | Kcl, HCTZ, Mg |
| GIT394-1 | - | - | - | - | - | F | - |
| GIT396-1 | - | - | - | - | - | M | - |
| GIT398-1 | 120/75 | 2.80 | 1.22 | 35.00 | 0.09 | M | Kcl, Mg, indocin |
| GIT411-1 | 130/92 | 3.10 | 1.00 | 29.00 | 0.01 | M | Mg, K, indocin |
| GIT418-1 | 120/80 | 3.20 | 1.56 | 31.10 | - | M | Kcl, Mg, indocin |
| GIT419-1 | 130/80 | 2.60 | 1.14 | 34.00 | - | M | Mg |
| GIT420-1 | 132/80 | 3.10 | 1.50 | 25.00 | 0.09 | F | Ca |
| GIT422-1 | 100/70 | 2.70 | 1.10 | 31.00 | 0.05 | F | KCl, Mg, spironolactone |
| GIT430-1 | 130/60 | 1.60 | 1.50 | 29.00 | 0.57 | F | K, Mg oxide, vitamin, birth |
| GIT431-1 | 106/70 | 2.60 | 1.20 | 28.00 | 0.05 | F | Mg, Kcl |
| GIT433-1 | 94/55 | 2.50 | 1.30 | 30.00 | 0.08 | F | KCl, |
| GIT435-1 | 112/70 | 3.90 | 1.70 | 23.00 | 0.35 | F | K |
| GIT438-1 | - | 3.00 | - | - | - | F | K, Mg |
| GIT439-1 | 101/57 | 4.80 | 2.28 | - | - | M | KCl |
| GIT447-1 | 98/62 | 2.60 | 1.56 | 28.00 | 0.03 | F | KCl |

**Table S1. List of 221 probands referred for genetic analyses.** Clinical information, demographics and medications are shown.

| PID | BP | K+ | Mg2+ | HCO3- | UCa2+/Cr | Gender | Medications |
|---|---|---|---|---|---|---|---|
| GIT452-1 | - | - | - | - | - | M | - |
| GIT453-1 | - | - | - | - | - | M | - |
| GIT459-1 | 118/70 | 3.30 | 1.10 | 25.00 | 0.10 | M | KCl |
| GIT460-1 | 120/78 | 3.00 | 1.50 | 31.00 | 1.00 | M | Kcl, Mg, Amiloride |
| GIT463-1 | - | - | - | - | - | M | - |
| GIT464-1 | - | - | - | - | - | F | - |
| GIT466-1 | - | - | - | - | - | F | - |
| GIT467-1 | - | - | - | - | - | M | - |
| GIT469-1 | 126/74 | 3.90 | 1.50 | 23.00 | 0.21 | F | Mg |
| GIT471-1 | 110/80 | 3.20 | 1.60 | 31.00 | 0.00 | F | Kcl, Mg, indocin |
| GIT473-1 | 94/57 | 4.90 | 1.80 | 22.00 | 2.30 | F | indocin |
| GIT478-1 | 107/64 | 2.60 | 0.80 | 28.00 | - | F | K, aldactone |
| GIT483-1 | - | - | - | - | - | M | - |
| GIT484-1 | 110/70 | 2.60 | 0.50 | 27.00 | 1.30 | F | Mg, Kdur |
| GIT485-1 | 78/- | 4.70 | 0.86 | 22.00 | 2.20 | F | indocin |
| GIT486-1 | 110/70 | 3.10 | 1.30 | 24.00 | 0.07 | M | Kcl, Mg, Amiloride |
| GIT487-1 | 117/58 | 3.00 | 0.80 | 28.00 | - | M | Mg |
| GIT488-1 | 96/60 | 2.90 | 2.20 | 30.50 | - | F | Kcl, Indocin |
| GIT490-1 | 100/20 | 2.80 | 1.82 | 30.00 | 0.18 | M | - |
| GIT491-1 | - | 4.60 | - | 27.00 | - | M | MgOx |
| GIT492-1 | - | - | - | - | - | F | - |
| GIT493-1 | 80/20 | 2.50 | 1.95 | 32.00 | 2.52 | M | - |
| GIT494-1 | - | 2.40 | - | 29.60 | 0.26 | M | - |
| GIT495-1 | 90/50 | 2.20 | 1.95 | 33.00 | 2.00 | M | Indocin, KCl |
| GIT496-1 | 90/45 | 2.60 | 1.63 | 28.00 | - | M | Kcl |
| GIT499-1 | - | 3.85 | 1.39 | 29.40 | - | M | Mg, Ca, phosphate |
| GIT505-1 | - | - | - | - | - | M | - |
| GIT508-1 | 134/80 | 2.00 | 1.00 | 31.00 | 0.54 | M | Kcl, Mg |
| GIT509-1 | - | - | - | - | - | M | - |
| GIT511-1 | - | - | - | - | - | M | - |
| GIT513-1 | 96/56 | 1.60 | 1.20 | 28.00 | 0.00 | M | Kcl, amiloride |
| GIT521-1 | 116/78 | 3.80 | 1.70 | 31.60 | - | M | Kcl |
| GIT523-1 | - | - | - | - | - | F | - |
| GIT524-1 | - | 2.60 | 1.40 | 25.00 | 0.04 | M | Kcl, MgOx |
| GIT526-1 | 90/50 | 2.40 | 1.30 | 30.00 | 0.16 | M | aldactone, K |
| GIT528-1 | 90/51 | 5.20 | 2.60 | 26.00 | - | F | KCl |
| GIT531-1 | 92/60 | 2.90 | 1.63 | 26.00 | 0.05 | M | KCl, Mg |
| GIT532-1 | - | 3.10 | 1.70 | 22.00 | 0.40 | M | Kcl, HCTZ, indocin |
| GIT533-1 | - | 2.55 | 1.60 | 26.50 | - | F | - |
| GIT536-1 | - | 3.17 | 2.43 | 34.50 | 0.39 | M | NaCl, KCl, spironolactone |
| GIT537-1 | 134/100 | 3.80 | 1.20 | - | - | F | K, Mg, amiloride |
| GIT538-1 | - | - | - | - | - | M | - |
| GIT539-1 | - | - | - | - | - | F | - |
| GIT540-1 | - | - | - | - | - | M | - |
| GIT542-1 | - | - | - | - | - | M | - |
| GIT543-1 | - | - | - | - | - | M | - |
| GIT544-1 | - | - | - | - | - | M | - |
| GIT545-1 | - | - | - | - | - | F | - |
| GIT546-1 | - | - | - | - | - | M | - |
| GIT548-1 | - | - | - | - | - | M | - |
| GIT550-1 | - | - | - | - | - | M | - |
| GIT551-1 | - | - | - | - | - | M | - |
| GIT553-1 | - | 2.80 | 2.30 | 32.00 | - | F | spironolactone, K |
| GIT554-1 | 130/76 | 3.00 | 1.90 | 32.20 | 0.80 | M | K, spironolactone |
| GIT555-1 | 90/60 | 1.80 | 1.10 | 25.10 | 0.03 | M | KCl |
| GIT560-1 | 102/70 | 2.00 | 1.40 | 35.00 | 0.00 | M | K |
| GIT561-1 | 116/63 | 2.00 | 1.60 | 28.00 | 0.15 | F | Indocin, Kdur, Mg |
| GIT563-1 | 107/90 | 2.90 | 1.20 | 28.00 | 0.70 | F | KCl |
| GIT565-1 | 104/54 | 3.20 | 1.10 | 29.00 | 0.05 | F | Mg, KCl |
| GIT566-1 | 91/60 | 3.20 | 1.40 | - | - | F | indocin, KCl |
| GIT569-1 | 112/65 | 3.30 | 2.20 | 33.00 | 1.00 | M | KCl, aldactone |
| GIT571-1 | - | - | - | - | - | F | - |
| GIT573-1 | 110/70 | 2.80 | 1.50 | 32.00 | 0.03 | F | Mg, KCl |
| GIT574-1 | 116/76 | 3.19 | 1.60 | 29.00 | 0.50 | F | amiloride, K |
| GIT576-1 | 134/66 | 3.50 | 1.30 | 27.00 | - | F | KCl |
| GIT578-1 | 130/80 | 2.80 | 2.00 | 38.00 | 0.30 | M | K, amiloride, KCl |
| GIT579-1 | 86/46 | 2.40 | - | 49.00 | 1.50 | F | - |
| GIT581-1 | - | - | - | - | - | M | indomethacin, KCl |
| GIT582-1 | - | - | - | - | - | F | aldactone, KCl |
| GIT584-1 | 130/80 | 3.50 | 2.30 | 21.00 | 0.03 | F | Mg |
| GIT585-1 | 100/75 | 2.40 | 1.56 | 31.00 | 0.08 | F | - |
| GIT588-1 | 120/80 | 4.50 | 2.00 | 26.00 | - | F | amiloride, K, Mg |

### Table S1 (cont.)

| PID | BP | K+ | Mg2+ | HCO3- | UCa2+/Cr | Gender | Medications |
|---|---|---|---|---|---|---|---|
| GIT589-1 | 130/80 | 2.64 | 1.97 | 28.00 | 0.05 | F | Kcl |
| GIT590-1 | 137/83 | 2.40 | 1.00 | 30.00 | 0.50 | F | Kcl, Mg, amiloride |
| GIT591-1 | 109/60 | 3.30 | 1.78 | 29.00 | - | F | - |
| GIT594-1 | - | - | - | - | - | M | - |
| GIT595-1 | 98/50 | 2.07 | 2.00 | 28.00 | - | F | KCl, spironolactone |
| GIT596-1 | 119/58 | 2.50 | 1.70 | 26.00 | - | F | aldactone, Kcl |
| GIT598-1 | - | - | - | - | - | F | amiloride, KCl, Mg |
| GIT601-1 | - | 4.10 | 1.10 | 29.00 | - | M | Mg, Amiloride |
| GIT602-1 | 103/60 | 2.10 | 2.20 | 30.00 | 0.72 | F | NaCl, KCl |
| GIT604-1 | 100/60 | 2.42 | 1.89 | 26.60 | 0.50 | M | Indocin, KCl |
| GIT605-1 | 90/60 | 2.50 | 1.70 | 59.90 | 0.54 | M | KCl, indocin |
| GIT606-1 | 100/65 | 2.30 | - | - | - | M | aldactone, KCl |
| GIT607-1 | - | - | - | - | - | F | - |
| GIT609-1 | - | 6.50 | 3.00 | 24.00 | - | M | KCl, spironolactone |
| GIT610-1 | - | 2.90 | 0.93 | 32.00 | - | F | - |
| GIT612-1 | - | - | - | - | - | M | - |
| GIT613-1 | 110/92 | 2.50 | 1.40 | 32.00 | - | M | Kcl, Mg, Amiloride |
| GIT614-1 | - | 2.90 | 1.30 | 30.00 | 0.50 | M | kcl, mg |
| GIT615-1 | - | 4.90 | 1.21 | 25.00 | - | M | K |
| GIT616-1 | 120/74 | 3.50 | 1.40 | 26.00 | - | F | Kcl |
| GIT617-1 | - | - | - | - | - | F | - |
| GIT618-1 | - | - | - | - | - | M | Kcl, Nacl |
| GIT619-1 | 118/58 | 2.30 | 2.60 | 32.10 | - | F | KCl, Spironolactone, [illegible] |
| GIT623-1 | - | - | - | - | - | F | - |
| GIT625-1 | - | - | - | - | - | M | - |
| GIT626-1 | 136/80 | 3.80 | 1.60 | 29.00 | - | M | Kdur, Mg |
| GIT627-1 | 134/80 | 2.30 | 1.10 | 34.00 | 0.88 | F | Kdur, MgOx |
| GIT628-1 | 110/75 | 2.40 | 0.90 | 31.00 | - | M | Kcl, MgOx |
| GIT629-1 | - | - | - | - | - | F | - |
| GIT631-1 | - | 2.60 | - | 38.00 | 1.00 | M | Kcl, indocin, NaCl |
| GIT633-1 | 120/70 | 2.40 | - | - | - | F | K |
| GIT634-1 | 130/78 | 1.80 | - | - | - | F | K |
| GIT636-1 | 120/80 | 2.40 | - | - | - | M | Kcl, indocin |
| GIT640-1 | 102/58 | 4.00 | 1.75 | 28.00 | - | F | K, Mg |
| GIT641-1 | 100/65 | 2.50 | 1.40 | 36.00 | 0.02 | M | Kcl, Mg |
| GIT642-1 | 130/64 | 2.30 | 1.30 | 27.00 | 0.08 | F | K, Mg, amiloride |
| GIT644-1 | - | - | - | - | - | M | - |
| GIT645-1 | 94/58 | 4.00 | - | - | - | M | - |
| GIT647-1 | - | - | - | - | - | M | - |
| GIT648-1 | 134/84 | 2.80 | 1.90 | 26.00 | - | F | Kcl |
| GIT649-1 | - | - | - | - | - | M | Kcl |
| GIT651-1 | - | - | - | - | - | F | - |
| GIT653-1 | 105/70 | 3.00 | - | - | - | M | Kdur, Mg |
| GIT654-1 | 108/84 | 4.20 | 1.90 | 23.90 | - | M | Kcl, Mg |
| GIT656-1 | 90/60 | 3.30 | - | - | - | M | Kcl |
| GIT659-1 | 95/57 | 2.65 | 1.15 | - | - | F | Mg |
| GIT670-1 | 110/98 | 3.40 | 1.00 | 29.00 | - | M | Mg, spironolactone |
| GIT671-1 | - | - | - | - | - | F | - |
| GIT677-1 | 74/52 | 2.10 | - | 30.00 | - | M | Kcl, Nacl, aldactone, |
| GIT679-1 | - | - | - | - | - | F | - |
| GIT681-1 | 100/62 | 2.80 | 1.80 | 29.00 | 0.23 | M | - |
| GIT684-1 | - | - | - | - | - | F | - |
| GIT689-1 | 111/56 | 2.60 | 1.80 | 24.00 | - | M | K |
| GIT691-1 | 90/54 | 1.90 | 1.30 | 29.00 | 0.18 | F | Kdur, Mg |
| GIT692-1 | 136/82 | 2.30 | 1.65 | 30.00 | - | F | Kcl |
| GIT695-1 | 88/55 | 3.10 | 1.70 | 30.00 | - | F | Kcl, Mg |
| GIT696-1 | - | 3.50 | 1.10 | - | - | F | Mg |
| GIT699-1 | 116/63 | 2.10 | 1.80 | 36.00 | 0.01 | M | Kcl, amiloride |
| GIT701-1 | - | 2.20 | 1.80 | 32.00 | - | M | Kcl, Nacl, aldactone, |
| GIT702-1 | 106/70 | 3.50 | 2.30 | 30.00 | - | M | spironolactone |
| GIT703-1 | 64/36 | 4.60 | 1.50 | 26.00 | - | F | Kcl, Nacl, indocin |
| GIT705-1 | 114/70 | 2.90 | 1.60 | 36.00 | - | M | K, Mg |
| GIT706-1 | 110/70 | 2.40 | - | 29.00 | 0.03 | F | K |
| GIT708-1 | - | 3.60 | 2.20 | - | - | F | K, Mg |
| GIT712-1 | 90/60 | 3.20 | 1.20 | - | - | F | Kcl, indocin |
| GIT719-1 | 100/60 | 2.40 | 2.10 | 33.00 | - | M | K |
| GIT723-1 | - | - | - | - | - | F | - |
| GIT725-1 | - | - | - | - | - | M | - |
| GIT727-1 | 121/69 | 2.90 | 1.30 | 28.00 | 0.05 | F | Kcl, Mg |
| GIT728-1 | 110/70 | 3.10 | 0.90 | 26.00 | 0.30 | F | Mg |
| GIT729-1 | - | 2.25 | - | - | - | M | Kcl, Mg, Nacl, amiloride |
| GIT731-1 | - | - | - | - | - | M | - |
| GIT734-1 | 104/53 | 2.60 | 1.90 | 35.00 | - | F | Kcl |
| GIT736-1 | - | - | - | - | - | M | Kcl, Mg, aldactone,, NaCl |
| GIT739-1 | 60/40 | 2.80 | 1.30 | 30.00 | - | M | Indocin |
| GIT741-1 | 90/39 | 2.20 | 1.60 | 28.00 | 0.03 | M | KCL, spironolactone |
| GIT741-2 | 100/51 | 2.60 | 1.70 | 28.00 | - | F | KCL, spironolactone |

**Table S1 (cont.)**

| Category | Probands sequenced with IDT | Probands sequenced with MedExome |
|---|---|---|
| Read length (bp) | 100.4 | 76.0 |
| # of reads per sample (M) | 65.3 | 74.4 |
| Median coverage at each targeted base (X) | 70.9 | 37.9 |
| Mean coverage at each targeted base (X) | 74.2 | 44.5 |
| % of all reads that map to tareget | 59.10% | 45.02% |
| % of all bases that map to target | 44.00% | 36.85% |
| % of targeted bases read at least 10X | 98.33% | 95.45% |
| % of targeted bases read at least 20X | 97.09% | 84.57% |
| % of targeted bases read at least 30X | 93.09% | 66.09% |
| % of targeted bases read at least 40X | 85.14% | 46.56% |
| % Mean error rate | 0.21% | 0.36% |

**Table S2. Whole-exome sequencing metrics for the salt-wasting probands performed by YCGA.**

| Category | Probands |
|---|---|
| Read length (bp) | 143.6 |
| # of reads per sample (M) | 912.4 |
| Median coverage at each targeted base (X) | 37.8 |
| Mean coverage at each targeted base (X) | 37.5 |
| % of all reads that map to target | 80.62% |
| % of all bases that map to target | 80.12% |
| % of targeted bases read at least 10X | 98.55% |
| % of targeted bases read at least 20X | 95.75% |
| % of targeted bases read at least 30X | 76.36% |
| % Mean error rate | 0.49% |

**Table S3. Whole genome sequencing metrics for the salt wasting probands with monoallelic SLC12A3 mutations.**

| PID | BP | $K^+$ | $Mg^{2+}$ | $HCO_3^-$ | $UCa^{2+}$:Cr | gene | genotype | mutation type | mutation | MetaSVM | CADD20 | gnomAD WES | ClinVar |
|---|---|---|---|---|---|---|---|---|---|---|---|---|---|
| GIT100-1 | 80/40 | 2.6 | 1.3 | 30 | - | SLC12A3 | homozygote | Dmis | L859P | D | 26.3 | 1.00E-04 | Pathogenic |
| GIT105-1 | 118/60 | 2.8 | 0.80 | 35 | 0.106 | SLC12A3 | homozygote | splicing | c.2883+1G>T | - | 34 | 2.80E-04 | Pathogenic |
| GIT107-1 | - | 2 | 1.30 | 33 | 0.16 | SLC12A3 | homozygote | splicing | c.1180+1G>T | - | 35 | 2.80E-05 | Pathogenic |
| GIT111-1 | - | 2.8 | 0.7 | 32 | 0.0001 | SLC12A3 | homozygote | splicing | c.1180+1G>T | - | 35 | 2.80E-05 | Pathogenic |
| GIT113-1 | 120/80 | 3.2 | 1.10 | 31 | 1.05 | SLC12A3 | homozygote | Dmis | R861C | D | 28 | 9.76E-05 | Pathogenic |
| GIT116-1 | 120/65 | 2.1 | 0.8 | 29 | 0.01 | SLC12A3 | homozygote | splicing | c.1180+1G>T | - | 35 | 2.80E-05 | Pathogenic |
| GIT117-1 | 110/60 | 2.1 | 1.2 | 32 | 0.09 | SLC12A3 | homozygote | splicing | c.2883+1G>T | - | 34 | 2.80E-04 | Pathogenic |
| GIT120-1 | 120/70 | 2.7 | 1.5 | 27 | 0.103 | SLC12A3 | homozygote | Dmis | G439S | D | 32 | 2.00E-04 | Pathogenic |
| GIT133-1 | 126/75 | 2.1 | 1 | 32 | 0.06 | SLC12A3 | homozygote | splicing | c.1180+1G>T | - | 35 | 2.80E-05 | Pathogenic |
| GIT141-1 | - | 2.80 | 1.50 | 37.00 | - | SLC12A3 | homozygote | Dmis | M343K | - | - | 0 | . |
| GIT143-1 | 90/50 | 2.5 | 1.90 | 25.6 | 0.01 | SLC12A3 | homozygote | Dmis | G741R | D | 27.5 | 0.0004 | Pathogenic |
| GIT187-1 | - | 2.4 | 1.33 | 27 | - | SLC12A3 | homozygote | splicing | c.1180+1G>T | - | 35 | 2.80E-05 | Pathogenic |
| GIT188-1 | - | 2.2 | 0.97 | 23.5 | 0.05 | SLC12A3 | homozygote | splicing | c.2883+1G>T | - | 34 | 2.80E-04 | Pathogenic |
| GIT194-1 | - | 2.1 | 1.20 | 30.5 | - | SLC12A3 | homozygote | Dmis | R1018X | - | 44 | 8.12E-06 | Pathogenic |
| GIT200-1 | - | 2.5 | 1.50 | 34 | 0.001 | SLC12A3 | homozygote | fs_ins | L998FS | - | - | 0 | . |
| GIT202-1 | 95/60 | 2.9 | 1.50 | - | - | SLC12A3 | homozygote | Dmis | V647M | - | 20.2 | 4.00E-05 | . |
| GIT204-1 | 109/73 | 2.5 | 1.10 | 29.9 | 0.11 | SLC12A3 | homozygote | splicing | c.1180+1G>T | - | 35 | 2.80E-05 | Pathogenic |
| GIT219-1 | - | 2.8 | 1.30 | 33.3 | 0.03 | SLC12A3 | homozygote | splicing | c.2883+1G>T | - | 34 | 2.80E-04 | Pathogenic |
| GIT222-1 | 116/82 | 2.5 | 1.30 | 35 | 0.04 | SLC12A3 | homozygote | splicing | c.2883+1G>T | - | 34 | 2.80E-04 | Pathogenic |
| GIT230-1 | 124/70 | 3.2 | 0.62 | 28 | - | SLC12A3 | homozygote | Dmis | D486N | D | 28.4 | 2.60E-05 | Pathogenic |
| GIT246-1 | - | 2.3 | 0.97 | 34.3 | 0.06 | SLC12A3 | homozygote | fs_ins | A401FS | - | - | 0 | . |
| GIT254-1 | 110/70 | 1.9 | 0.90 | 32 | - | SLC12A3 | homozygote | splicing | c.1568-1G>A | - | 33 | 8.00E-06 | Pathogenic |
| GIT269-1 | 100/60 | 2.2 | 1.43 | 25.7 | 0.56 | SLC12A3 | homozygote | splicing | c.1180+1G>T | - | 35 | 2.80E-05 | Pathogenic |
| GIT300-1 | 122/69 | 2.9 | 1 | 26.6 | 0.11 | SLC12A3 | homozygote | Dmis | G741R | D | 27.5 | 4.00E-04 | Pathogenic |
| GIT301-1 | - | 2.70 | 1.17 | - | 0.05 | SLC12A3 | homozygote | splicing | c.1180+1G>T | - | 26.4 | 3.00E-05 | . |
| GIT317-1 | - | 3 | 1.10 | 32 | - | SLC12A3 | homozygote | stopgain | K497X | - | 43 | 1.75E-05 | Pathogenic |
| GIT322-1 | - | 2.70 | 0.80 | 28.10 | - | SLC12A3 | homozygote | Dmis | G741R | D | 27.5 | 4.00E-04 | Pathogenic |
| GIT328-1 | 110/80 | 2.70 | 1.20 | 27.00 | 0.03 | SLC12A3 | homozygote | CNV | ex9del | - | - | 0.00E+00 | . |
| GIT367-1 | 100/70 | 2.4 | 1.70 | 30 | 0.06 | SLC12A3 | homozygote | fs_del | T697FS | - | 34 | 2.50E-05 | Pathogenic |
| GIT368-1 | 110/85 | 2.7 | 1.40 | 29.4 | 0.01 | SLC12A3 | homozygote | Dmis | S178L | - | 25.1 | 1.60E-05 | Pathogenic |
| GIT407-1 | - | - | - | - | - | SLC12A3 | homozygote | fs_del | L15FS | - | - | 0.00E+00 | Pathogenic |
| GIT432-1 | - | 1.90 | 1.00 | - | - | SLC12A3 | homozygote | Dmis | L246V | - | - | 0.00E+00 | . |
| GIT436-1 | - | 2.60 | - | 29.80 | - | SLC12A3 | homozygote | Dmis | P643L | D | 34 | 2.00E-04 | Pathogenic |
| GIT471-1 | 110/80 | 3.20 | 1.60 | 31.00 | 0.00 | SLC12A3 | homozygote | Dmis | R399C | D | 28.7 | 1.20E-05 | Pathogenic |
| GIT508-1 | 134/80 | 2.00 | 1.00 | 31.00 | 0.54 | SLC12A3 | homozygote | splicing | c.506-1G>A | - | 24.4 | 4.40E-05 | . |
| GIT533-1 | - | 2.55 | 1.60 | 26.50 | - | SLC12A3 | homozygote | Dmis | P643L | D | 34 | 2.00E-04 | Pathogenic |
| GIT555-1 | 90/60 | 1.80 | 1.10 | 25.10 | 0.03 | SLC12A3 | homozygote | Dmis | G439S | D | 32 | 2.00E-04 | Pathogenic |
| GIT560-1 | 102/70 | 2.00 | 1.40 | 35.00 | 0.00 | SLC12A3 | homozygote | Dmis | T649M | D | 32 | 3.70E-05 | Pathogenic |
| GIT585-1 | 100/75 | 2.40 | 1.56 | 31.00 | 0.08 | SLC12A3 | homozygote | Dmis | L859P | D | 26.3 | 1.00E-04 | Pathogenic |
| GIT590-1 | 137/83 | 2.40 | 1.00 | 30.00 | 0.50 | SLC12A3 | homozygote | Dmis | G741R | D | 27.5 | 4.00E-04 | Pathogenic |
| GIT613-1 | 110/92 | 2.50 | 1.40 | 32.00 | - | SLC12A3 | homozygote | Dmis | L859P | D | 26.3 | 1.00E-04 | Pathogenic |
| GIT625-1 | - | - | - | - | - | SLC12A3 | homozygote | splicing | c.602-16G>A | . | . | 6.00E-05 | - |
| GIT628-1 | 110/75 | 2.40 | 0.90 | 31.00 | - | SLC12A3 | homozygote | Dmis | R334Q | T | 29.3 | 1.20E-05 | . |
| GIT659-1 | 95/57 | 2.65 | 1.15 | - | - | SLC12A3 | homozygote | stopgain | W746X | - | 42 | 0.00E+00 | . |
| GIT681-1 | 100/62 | 2.80 | 1.80 | 29.00 | 0.23 | SLC12A3 | homozygote | Dmis | P643L | D | 34 | 2.00E-04 | Pathogenic |
| GIT719-1 | 100/60 | 2.40 | 2.10 | 33.00 | - | SLC12A3 | homozygote | Dmis | G729A | D | 25.6 | 0.00E+00 | . |

**Table S4. List of probands with homozygous mutations in SLC12A3.** Clinical information, followed by information on the biallelic mutation, is shown.

| PID | BP | $K^+$ | $Mg^{2+}$ | $HCO_3^-$ | $UCa^{2+}$:Cr | gene | genotype | mutation #1 type | mutation #1 | Meta SVM | CADD20 | gnomAD WES | ClinVar | mutation #2 type | mutation #2 | Meta SVM | CADD20 | gnomAD WES | ClinVar |
|---|---|---|---|---|---|---|---|---|---|---|---|---|---|---|---|---|---|---|---|
| GIT102-1 | 114/70 | 1.9 | 1.30 | 31.9 | - | SLC12A3 | compound het | Dmis | C421R | - | 28 | 0 | Pathogenic | Dmis | R209W | | | | Pathogenic |
| GIT103-1 | 110/60 | 2.5 | 1.20 | 36 | - | SLC12A3 | compound het | Dmis | K284R | - | 32 | 0.00E+00 | . | Dmis | R655H | - | 29 | 0.00008499 | Pathogenic |
| GIT104-1 | 110/70 | 1 | 1.10 | 30 | 0.24 | SLC12A3 | compound het | splicing | c.2883+1G>T | - | 34 | 2.80E-04 | Pathogenic | Dmis | R655L | - | 26.6 | 8.00E-06 | Pathogenic |
| GIT106-1 | - | 2.20 | 1.30 | 33.00 | - | SLC12A3 | compound het | Dmis | A588V | - | 27.5 | 2.83E-05 | Pathogenic | Dmis | L859P | D | 26.3 | 0.0001 | Pathogenic |
| GIT108-1 | 130/80 | 2.2 | 1.5 | 22 | 0.08 | SLC12A3 | compound het | Dmis | G630V | - | - | 0 | Pathogenic | nfs_del | Del S561 | - | - | 0 | . |
| GIT110-1 | 110/60 | 3.00 | 1.22 | 31.00 | 0.08 | SLC12A3 | compound het | splicing | c.2856+1G>T | - | 24.3 | 2.00E-04 | . | Dmis | G496C | D | 34 | 2.20E-05 | . |
| GIT112-1 | - | 2.5 | 1.20 | 30 | 0.13 | SLC12A3 | compound het | Dmis | G741R | D | 27.5 | 0.0004 | Pathogenic | stopgain | R977X | - | 49 | 8.00E-06 | Pathogenic |
| GIT114-1 | 100/60 | 2.6 | 1.10 | 29 | 0.20 | SLC12A3 | compound het | Dmis | C994Y | D | 25.7 | 1.90E-04 | . | splicing | c.2883+1G>T | - | 34 | 0.000028 | Pathogenic |
| GIT115-1 | 120/70 | 2.8 | 1.10 | 33.2 | - | SLC12A3 | compound het | Dmis | G741R | D | 27.5 | 0.0004 | Pathogenic | splicing | c.2883+1G>T | - | 34 | 0.000028 | Pathogenic |
| GIT118-1 | 140/80 | 2.2 | 1.6 | 32 | 0.056 | SLC12A3 | compound het | Dmis | G439S | D | 32 | 2.00E-04 | Pathogenic | Tmis | H69N | - | 17.53 | 4.00E-06 | . |
| GIT119-1 | 140/90 | 2.8 | 1.5 | 30 | 0.042 | SLC12A3 | compound het | splicing | c.2883+1G>T | - | 34 | 2.80E-04 | Pathogenic | Dmis | R964Q | - | 29.3 | 1.50E-04 | Pathogenic |
| GIT121-1 | 120/80 | 2.5 | 1.2 | 30 | 0.072 | SLC12A3 | compound het | Dmis | K734N | - | - | 0.00E+00 | . | Dmis | R964Q | - | 29.3 | 1.50E-04 | Pathogenic |
| GIT122-1 | 140/85 | 2.5 | 1.2 | 26 | 0.043 | SLC12A3 | compound het | fs_del | T697FS | - | 34 | 2.50E-05 | Pathogenic | Dmis | G729V | D | 27.6 | 0.0001 | Pathogenic |
| GIT123-1 | 120/60 | 2.60 | 1.27 | 32.00 | 0.01 | SLC12A3 | compound het | Dmis | R334W | D | 24.8 | 2.10E-05 | . | CNV | ex2-3 del | - | - | 0 | . |
| GIT124-1 | 120/80 | 2.7 | 0.9 | 37 | 0.11 | SLC12A3 | compound het | Dmis | G439S | D | 32 | 2.00E-04 | Pathogenic | fs_del | G731FS | - | - | 0 | . |
| GIT126-1 | 130/65 | 2.7 | 1.20 | 28 | 0.01 | SLC12A3 | compound het | Dmis | R334W | D | 24.8 | 2.10E-05 | . | Dmis | R399C | - | 25.5 | 1.20E-05 | Pathogenic |
| GIT132-1 | - | 2.80 | 1.30 | - | 0.18 | SLC12A3 | compound het | Dmis | G741R | D | 27.5 | 0.0004 | Pathogenic | Dmis | R642H | D | 35 | 0 | Pathogenic |
| GIT134-1 | - | 2.50 | 1.20 | 31.00 | 0.00 | SLC12A3 | compound het | fs_ins | A401FS | - | - | 0 | . | Dmis | G460D | - | - | 0.00E+00 | . |
| GIT135-1 | - | 2.20 | 1.30 | 28.60 | - | SLC12A3 | compound het | splicing | c.2883+1G>T | - | 34 | 2.80E-04 | Pathogenic | Dmis | R399L | - | 26.4 | 2.90E-05 | Pathogenic |
| GIT136-1 | - | - | - | - | - | SLC12A3 | compound het | Dmis | R321W | - | 23.9 | 8.90E-05 | Pathogenic | Dmis | L859P | D | 26.3 | 0.0001 | Pathogenic |
| GIT137-1 | 108/64 | 3 | 1.10 | 26.3 | - | SLC12A3 | compound het | Dmis | R209Q | - | 32 | 2.83E-05 | Pathogenic | stopgain | K497X | - | 43 | 1.75E-05 | Pathogenic |
| GIT140-1 | 130/70 | 2.5 | 1.3 | 32 | 0.19 | SLC12A3 | compound het | CNV | del (1-7) exons | - | - | 0 | . | Dmis | R642G | D | 33 | 2.50E-05 | Pathogenic |
| GIT146-1 | - | 2.70 | 0.90 | 32.90 | 0.13 | SLC12A3 | compound het | Dmis | M581K | - | 26.8 | 7.40E-05 | Pathogenic | Dmis | V1024M | - | 29.6 | 1.20E-05 | . |
| GIT150-1 | 80/50 | 2.10 | 1.70 | 30.00 | 0.34 | SLC12A3 | compound het | Dmis | G741R | D | 27.5 | 4.00E-04 | Pathogenic | CNV | ex2-3 del | - | - | 0 | . |
| GIT151-1 | 80/60 | 2.80 | 1.68 | - | 0.31 | SLC12A3 | compound het | CNV | ex2-3 del | - | - | 0.00E+00 | . | Dmis | G741R | D | 27.5 | 0.0004 | Pathogenic |
| GIT153-1 | - | 3.10 | 1.10 | 34.00 | - | SLC12A3 | compound het | Dmis | D486N | D | 28.4 | 2.60E-05 | Pathogenic | Dmis | R964Q | - | 29.3 | 1.50E-04 | Pathogenic |
| GIT158-1 | 96/60 | 3.5 | 1.1 | 32 | 0.056 | SLC12A3 | compound het | Dmis | P643L | D | 34 | 2.00E-04 | Pathogenic | Dmis | R964Q | - | 29.3 | 1.50E-04 | Pathogenic |
| GIT163-1 | 116/68 | 2.4 | 1.60 | 27 | 0.25 | SLC12A3 | compound het | Dmis | R399L | - | 26.4 | 2.90E-05 | Pathogenic | Dmis | L859P | D | 26.3 | 0.0001 | Pathogenic |
| GIT166-1 | 94/58 | 2.6 | 1.40 | - | 0.48 | SLC12A3 | compound het | Dmis | R655H | - | 29 | 0.00008499 | Pathogenic | splicing | c.741+1G>A | - | - | 8.00E-06 | Pathogenic |
| GIT168-1 | - | 3.5 | 1.50 | 34.9 | - | SLC12A3 | compound het | splicing | c.2883+1G>T | - | 34 | 2.80E-04 | Pathogenic | Dmis | P643L | D | 34 | 2.00E-04 | Pathogenic |
| GIT170-1 | 130/80 | 3.1 | - | 34 | - | SLC12A3 | compound het | splicing | c.2883+1G>T | - | 34 | 2.80E-04 | Pathogenic | Dmis | G989R | T | 25.9 | 5.29E-05 | Pathogenic |
| GIT172-1 | 100/60 | 2.80 | 0.32 | 29.00 | 0.22 | SLC12A3 | compound het | Dmis | N442K | T | 26.4 | 4.00E-06 | Pathogenic | Dmis | S620P | T | 27 | 0 | . |
| GIT177-1 | - | 2.90 | 0.85 | 30.00 | 0.11 | SLC12A3 | compound het | stopgain | E112X | - | 42 | 1.10E-05 | Pathogenic | Dmis | G989R | T | 25.9 | 5.29E-05 | Pathogenic |
| GIT190-1 | - | 2.7 | 0.9 | 27 | - | SLC12A3 | compound het | Dmis | R861C | D | 28 | 9.76E-05 | Pathogenic | Dmis | R896Q | - | 32 | 3.50E-05 | Pathogenic |
| GIT192-1 | - | - | - | - | - | SLC12A3 | compound het | Dmis | R852C | D | 28 | 9.76E-05 | Pathogenic | branch A del | c.2447-37_2447-19del | - | - | 1.50E-02 | . |
| GIT193-1 | 120/80 | 2.80 | 0.80 | 30.00 | 0.00 | SLC12A3 | compound het | Dmis | R158Q | D | 34 | 4.10E-05 | Pathogenic | fs_del | T697fs | - | - | 2.84E-05 | Pathogenic |
| GIT195-1 | - | 2.5 | 1.50 | 34 | 0.14 | SLC12A3 | compound het | fs_del | T697FS | - | 34 | 2.50E-05 | Pathogenic | Dmis | R871P | - | - | 0 | Pathogenic |
| GIT199-1 | 106/76 | 2.5 | 1.00 | 36 | 0.05 | SLC12A3 | compound het | splicing | c.2883+1G>T | - | 34 | 2.80E-04 | Pathogenic | splicing | c.2883+1G>T | - | 34 | 0.000028 | Pathogenic |
| GIT201-1 | - | 2.10 | 1.70 | 37.40 | - | SLC12A3 | compound het | Dmis | G439S | D | 32 | 2.00E-04 | Pathogenic | Dmis | L215P | T | 24.7 | 8.10E-06 | Pathogenic |
| GIT203-1 | - | 2.7 | 1.20 | 29 | - | SLC12A3 | compound het | Dmis | S555L | - | 27.6 | 6.00E-05 | Pathogenic | Dmis | L859P | D | 26.3 | 0.0001 | Pathogenic |

**Table S5. List of probands with compound heterozygous mutations in the salt-wasting cohort.** Clinical information, followed by information on the biallelic mutations, is shown.

| PID | BP | $K^+$ | $Mg^{2+}$ | $HCO_3^-$ | $UCa^{2+}$:Cr | gene | genotype | mutation #1 type | mutation #1 | Meta SVM | CADD20 | gnomAD WES | ClinVar | mutation #2 type | mutation #2 | Meta SVM | CADD20 | gnomAD WES | ClinVar |
|---|---|---|---|---|---|---|---|---|---|---|---|---|---|---|---|---|---|---|---|
| GIT212-1 | 108/62 | 2.4 | 1.20 | 37 | 0.34 | SLC12A3 | compound het | Dmis | L859P | D | 26.3 | 1.00E-04 | Pathogenic | Dmis | T1026I | - | 26.2 | 8.00E-06 | Pathogenic |
| GIT213-1 | - | 2.7 | 1.40 | 30 | 0.22 | SLC12A3 | compound het | Dmis | G741R | D | 27.5 | 4.00E-04 | Pathogenic | splicing | c.1180+1G>T | - | 35 | 2.80E-05 | Pathogenic |
| GIT214-1 | 110/60 | 2.30 | 1.79 | 30.00 | - | SLC12A3 | compound het | Dmis | G741R | D | 27.5 | 4.00E-04 | Pathogenic | Dmis | G439S | D | 32 | 2.00E-04 | Pathogenic |
| GIT216-1 | 100/80 | 1.3 | 1.10 | 31 | 0.82 | SLC12A3 | compound het | Dmis | R642H | - | 35 | 0.00E+00 | Pathogenic | Dmis | L1010R | - | - | 0.00E+00 | . |
| GIT217-1 | - | - | - | - | - | SLC12A3 | compound het | fs_ins | E78fs | - | - | 0.00E+00 | . | Dmis | A464T | T | 32 | 1.20E-05 | Pathogenic |
| GIT224-1 | - | 2.90 | 0.9 | 26.90 | - | SLC12A3 | compound het | Dmis | A459D | - | 25.3 | 3.20E-05 | . | Dmis | G989R | T | 25.9 | 5.29E-05 | Pathogenic |
| GIT228-1 | 104/74 | 3 | 1.6 | 30 | - | SLC12A3 | compound het | fs_del | W948FS | - | - | 0.00E+00 | Pathogenic | splicing | c.2883+1G>T | - | 34 | 2.80E-04 | Pathogenic |
| GIT229-1 | 104/40 | 2.6 | 1.20 | 32 | 0.05 | SLC12A3 | compound het | Dmis | R321W | - | 23.9 | 8.90E-05 | Pathogenic | splicing | c.2883+1G>T | - | 34 | 2.80E-04 | Pathogenic |
| GIT237-1 | 140/70 | 3.3 | 1.06 | 35 | - | SLC12A3 | compound het | Dmis | I988T | - | - | 0.00E+00 | . | splicing | c.2883+1G>T | - | 34 | 0.000028 | Pathogenic |
| GIT240-1 | - | - | - | - | - | SLC12A3 | compound het | Dmis | G741R | D | 27.5 | 4.00E-04 | Pathogenic | Dmis | M143V | T | 22.6 | 0 | . |
| GIT241-1 | 150/100 | 2.5 | 1.8 | 30 | - | SLC12A3 | compound het | fs_ins | A401FS | - | - | 0 | . | Dmis | R145C | | | | . |
| GIT245-1 | 110/70 | 2.6 | 1.1 | 29 | - | SLC12A3 | compound het | Dmis | S402Y | - | 32 | 0 | . | Dmis | R642G | D | 33 | 2.50E-05 | Pathogenic |
| GIT248-1 | - | 2.80 | 1.40 | 29.00 | - | SLC12A3 | compound het | Dmis | G439S | D | 32 | 2.00E-04 | Pathogenic | splicing | c.602-2A>G | - | - | 0 | . |
| GIT253-1 | 128/96 | 3 | 1.00 | - | 0.15 | SLC12A3 | compound het | splicing | c.2883+1G>T | - | 34 | 2.80E-04 | Pathogenic | splicing | c.1670-191 GC>GT | - | - | 3.2E-05 | . |
| GIT260-1 | 110/60 | 2.7 | 1.80 | 29 | 0.05 | SLC12A3 | compound het | Dmis | I904F | - | - | 0.00E+00 | . | splicing | c.1180+1G>T | - | 35 | 2.80E-05 | Pathogenic |
| GIT261-1 | - | 2.9 | 1.40 | 34 | - | SLC12A3 | compound het | Dmis | G741R | D | 27.5 | 4.00E-04 | Pathogenic | fs_del | T697FS | - | - | 2.84E-05 | Pathogenic |
| GIT262-1 | 125/78 | 2.9 | 0.92 | 29 | 0.00 | SLC12A3 | compound het | fs_ins | P79FS | - | - | 0 | . | Dmis | G741R | D | 27.5 | 0.0004 | Pathogenic |
| GIT271-1 | 141/82 | 3.30 | 0.80 | 27.00 | 0.00 | SLC12A3 | compound het | Dmis | A313V | T | 23 | 2.03E-05 | Pathogenic | Dmis | R887Q | D | 31 | 3.60E-05 | Pathogenic |
| GIT275-1 | 138/76 | 3.50 | 1.99 | 29.00 | 0.06 | SLC12A3 | compound het | fs_ins | E78fs | - | - | 0.00E+00 | . | stopgain | Y444X | - | 37 | 0 | Pathogenic |
| GIT280-1 | - | 2.90 | 1.44 | - | 0.00 | SLC12A3 | compound het | Dmis | A224D | - | 24.6 | 0 | . | Dmis | L738R | D | 29.4 | 0 | Pathogenic |
| GIT285-1 | 90/60 | 1.9 | 0.80 | 29 | 0.01 | SLC12A3 | compound het | Dmis | A313V | T | 23 | 2.03E-05 | Pathogenic | Dmis | G729V | D | 27.6 | 0.0001 | Pathogenic |
| GIT288-1 | - | 3 | 1.45 | 31.5 | 0.04 | SLC12A3 | compound het | Dmis | G729V | D | 27.6 | 0.0001 | Pathogenic | splicing | c.2883+1G>T | - | 34 | 0.000028 | Pathogenic |
| GIT289-1 | - | - | - | - | - | SLC12A3 | compound het | Dmis | G741R | D | 27.5 | 4.00E-04 | Pathogenic | Dmis | C994Y | D | 25.8 | 0.0002 | . |
| GIT293-1 | 118/74 | 3.00 | 1.90 | 33.00 | 0.07 | SLC12A3 | compound het | Dmis | S615W | D | 34 | 0 | Pathogenic | Dmis | G731R | D | 32 | 2.03E-05 | . |
| GIT295-1 | - | - | - | - | - | SLC12A3 | compound het | Dmis | R642C | D | 35 | 0.0001 | Pathogenic | fs_del | W939fs | - | - | 0 | Pathogenic |
| GIT297-1 | - | 3.5 | 1.2 | 25.1 | - | SLC12A3 | compound het | Dmis | A356V | - | 27.3 | 7.10E-06 | . | Dmis | G439S | D | 32 | 2.00E-04 | Pathogenic |
| GIT302-1 | 98/68 | 2.9 | 1.50 | 27 | - | SLC12A3 | compound het | fs_ins | A401FS | - | - | 0 | . | Dmis | G741R | D | 27.5 | 0.0004 | Pathogenic |
| GIT305-1 | 148/96 | 2.6 | 1.41 | 31 | 0.16 | SLC12A3 | compound het | splicing | c.2883+1G>T | - | 34 | 2.80E-04 | Pathogenic | Dmis | R321W | - | 23.9 | 8.90E-05 | . |
| GIT308-1 | - | 3.4 | 1.30 | 32 | 0.02 | SLC12A3 | compound het | Dmis | L272P | - | 26.3 | 1.80E-05 | Pathogenic | Dmis | G741R | D | 27.5 | 0.0004 | Pathogenic |
| GIT309-1 | 100/60 | 2.8 | 1.20 | - | 0.06 | SLC12A3 | compound het | Dmis | S555L | - | 27.6 | 6.00E-05 | Pathogenic | splicing | c.1568-1G>A | - | 34 | 2.00E-04 | Pathogenic |
| GIT314-1 | - | 3.70 | 1.33 | 30.00 | - | SLC12A3 | compound het | Dmis | R399C | - | 25.5 | 1.20E-05 | Pathogenic | Dmis | G613S | - | 29 | 3.20E-05 | . |
| GIT316-1 | 105/70 | 2.8 | 1.43 | 34 | 0.03 | SLC12A3 | compound het | Dmis | S178L | - | 25.1 | 1.60E-05 | Pathogenic | Dmis | P643L | D | 34 | 2.00E-04 | Pathogenic |
| GIT320-1 | 114/56 | 2.6 | 1.29 | 28 | - | SLC12A3 | compound het | Dmis | P735L | - | 25 | 0.00E+00 | . | Dmis | L859P | D | 26.3 | 0.0001 | Pathogenic |
| GIT330-1 | 100/55 | 2.9 | 1.6 | 26.8 | 0.40 | SLC12A3 | compound het | Dmis | T1026I | - | 26.2 | 8.00E-06 | Pathogenic | splicing | c.2883+1G>T | - | 34 | 0.000028 | Pathogenic |
| GIT337-1 | 120/60 | 2.9 | 1.32 | 33 | 0.13 | SLC12A3 | compound het | Dmis | G741R | D | 27.5 | 4.00E-04 | Pathogenic | Dmis | L859P | D | 26.3 | 0.0001 | Pathogenic |
| GIT341-1 | 105/80 | 2.5 | 0.90 | - | 0.07 | SLC12A3 | compound het | Dmis | I154F | - | 28.7 | 1.80E-05 | Pathogenic | Dmis | G741R | D | 27.5 | 0.0004 | Pathogenic |
| GIT348-1 | 110/60 | 2.90 | 1.41 | 31.00 | 0.06 | SLC12A3 | compound het | Dmis | L859P | D | 26.3 | 0.0001 | Pathogenic | stopgain | R1009X | - | 44 | 8.12E-06 | Pathogenic |
| GIT350-1 | 119/54 | 2.9 | 1.1 | 29 | 0.25 | SLC12A3 | compound het | Dmis | G439S | D | 32 | 2.00E-04 | Pathogenic | Dmis | Q910H | | | | . |
| GIT352-1 | 110/60 | 1.49 | 2.43 | 28 | 0.17 | SLC12A3 | compound het | Dmis | I192S | - | 28 | 4.00E-06 | . | Dmis | K706T | - | - | 0.00E+00 | . |
| GIT360-1 | 110/80 | 3.30 | 1.50 | 32.00 | 0.60 | SLC12A3 | compound het | Dmis | R642G | D | 33 | 2.64E-05 | Pathogenic | Dmis | R334W | D | 33 | 2.03E-05 | . |

### Table S5 (cont.)

| PID | BP | $K^+$ | $Mg^{2+}$ | $HCO_3^-$ | $UCa^{2+}$:Cr | gene | genotype | mutation #1 type | mutation #1 | Meta SVM | CADD20 | gnomAD WES | ClinVar | mutation #2 type | mutation #2 | Meta SVM | CADD20 | gnomAD WES | ClinVar |
|---|---|---|---|---|---|---|---|---|---|---|---|---|---|---|---|---|---|---|---|
| GIT361-1 | 118/66 | 2.4 | 1.70 | 32 | 0.17 | SLC12A3 | compound het | Dmis | R1018Q | D | 29 | 2.10E-05 | Pathogenic | Dmis | P643L | D | 34 | 2.00E-04 | Pathogenic |
| GIT363-1 | - | - | - | - | - | SLC12A3 | compound het | Dmis | R399C | - | 25.5 | 1.20E-05 | Pathogenic | Dmis | C994Y | D | 25.8 | 0.0002 | . |
| GIT364-1 | - | 3 | 1.00 | 31 | 0.17 | SLC12A3 | compound het | Dmis | G741R | D | 27.5 | 4.00E-04 | Pathogenic | splicing | c.2883+1G>T | - | 34 | 2.80E-04 | Pathogenic |
| GIT365-1 | 116/65 | 2.4 | 1.00 | 31 | 0.20 | SLC12A3 | compound het | Dmis | G741R | D | 27.5 | 4.00E-04 | Pathogenic | Dmis | L859P | D | 26.3 | 0.0001 | Pathogenic |
| GIT369-1 | 120/90 | 2.50 | 1.14 | 29.00 | 0.01 | SLC12A3 | compound het | Dmis | R158P | D | 34 | 0 | . | Dmis | S615L | D | 35 | 4.59E-05 | Pathogenic |
| GIT377-1 | 128/78 | 3.20 | 0.90 | 31.00 | 0.05 | SLC12A3 | compound het | Dmis | L859P | D | 26.3 | 0.0001 | Pathogenic | Dmis | C994Y | D | 25.8 | 0.0002 | . |
| GIT378-1 | 110/75 | 3.4 | 0.47 | 31 | - | SLC12A3 | compound het | Dmis | P643L | D | 34 | 2.00E-04 | Pathogenic | Dmis | C985Y | D | 25.7 | 0 | Pathogenic |
| GIT379-1 | 120/70 | 3.10 | 1.14 | - | 0.15 | SLC12A3 | compound het | stopgain | R1009X | - | 44 | 8.12E-06 | Pathogenic | Dmis | R83W | D | 34 | 2.85E-05 | Pathogenic |
| GIT384-1 | 120/78 | 2.80 | 1.70 | 27.00 | 0.06 | SLC12A3 | compound het | splicing | c.506-1G>A | - | 24.4 | 4.47E-05 | . | Dmis | R955W | T | 27.7 | 4.06E-05 | Pathogenic |
| GIT387-1 | 143/73 | 2.40 | 1.00 | 31.00 | 0.00 | SLC12A3 | compound het | Dmis | G741R | D | 27.5 | 4.00E-04 | Pathogenic | CNV | ex1-5 del | - | - | 0 | . |
| GIT388-1 | - | - | - | - | 2.33 | SLC12A3 | compound het | Dmis | L272P | - | 26.3 | 1.80E-05 | Pathogenic | Dmis | G989R | T | 25.9 | 5.29E-05 | Pathogenic |
| GIT391-1 | 130/80 | 2.4 | 1.9 | 31 | 0.17 | SLC12A3 | compound het | Dmis | G741R | D | 27.5 | 4.00E-04 | Pathogenic | splicing | c.2883+1G>T | - | 34 | 0.000028 | Pathogenic |
| GIT392-1 | 110/70 | 3.00 | 1.22 | 27.00 | - | SLC12A3 | compound het | Dmis | Y990C | D | 25.4 | 8.12E-06 | . | Dmis | Q1021R | T | 27 | 4.06E-06 | . |
| GIT411-1 | 130/92 | 3.10 | 1.00 | 29.00 | 0.01 | SLC12A3 | compound het | Dmis | G418R | T | 27.2 | 0 | . | Dmis | A523T | D | 29.6 | 1.09E-05 | Pathogenic |
| GIT413-1 | 110/60 | 2.04 | 2.30 | 31.00 | 0.15 | SLC12A3 | compound het | Dmis | T382M | D | 34 | 5.32E-05 | Pathogenic | Dmis | G741R | D | 27.5 | 0.0004 | Pathogenic |
| GIT416-1 | 110/70 | 2.9 | 1.4 | 31 | 0.09 | SLC12A3 | compound het | Dmis | T382M | D | 34 | 5.32E-05 | Pathogenic | splicing | c.2883+1G>T | - | 34 | 0.000028 | Pathogenic |
| GIT417-1 | 130/78 | 3.9 | 1.4 | 28.00 | 0.59 | SLC12A3 | compound het | Dmis | A166V | - | 33 | 3.20E-05 | . | Dmis | G439S | D | 32 | 2.00E-04 | Pathogenic |
| GIT419-1 | 130/80 | 2.60 | 1.14 | 34.00 | - | SLC12A3 | compound het | Dmis | G741R | D | 27.5 | 4.00E-04 | Pathogenic | Dmis | L859P | D | 26.3 | 0.0001 | Pathogenic |
| GIT422-1 | 100/70 | 2.70 | 1.10 | 31.00 | 0.05 | SLC12A3 | compound het | splicing | c.1180+1G>T | - | 35 | 2.80E-05 | Pathogenic | Dmis | R158L | D | 34 | . | . |
| GIT427-1 | - | 2.30 | 0.97 | 35.00 | - | SLC12A3 | compound het | Dmis | G439S | D | 32 | 2.00E-04 | Pathogenic | splicing | c.2548+1G>T | - | - | - | . |
| GIT430-1 | 130/60 | 1.60 | 1.50 | 29.00 | 0.57 | SLC12A3 | compound het | Dmis | G741R | D | 27.5 | 4.00E-04 | Pathogenic | splicing | c.2856+1G>T | - | 24.3 | 0.0002 | . |
| GIT431-1 | 106/70 | 2.60 | 1.20 | 28.00 | 0.05 | SLC12A3 | compound het | Dmis | G439S | D | 32 | 0.0002 | Pathogenic | fs_del | T7fs | - | - | 4.09E-06 | Pathogenic |
| GIT433-1 | 94/55 | 2.50 | 1.30 | 30.00 | 0.08 | SLC12A3 | compound het | splicing | c.2856+1G>T | - | 24.3 | 0.0002 | . | Dmis | G729V | D | 27.6 | 0.0001 | Pathogenic |
| GIT438-1 | - | 3.00 | - | - | - | SLC12A3 | compound het | Dmis | G741R | D | 27.5 | 4.00E-04 | Pathogenic | CNV | ex1-5 del | - | - | 0 | . |
| GIT445-1 | - | 3.50 | 1.60 | 29.00 | - | SLC12A3 | compound het | Dmis | G989R | T | 25.9 | 5.29E-05 | Pathogenic | fs_ins | F994FS | - | - | 0 | . |
| GIT447-1 | 98/62 | 2.60 | 1.56 | 28.00 | 0.03 | SLC12A3 | compound het | Dmis | L858H | D | 27.4 | 6.51E-05 | Pathogenic | fs_del | M991fs | - | - | 0 | . |
| GIT452-1 | - | - | - | - | - | SLC12A3 | compound het | splicing | c.1180+1G>T | - | 35 | 2.80E-05 | Pathogenic | Dmis | R642C | D | 35 | 0.0001 | Pathogenic |
| GIT459-1 | 118/70 | 3.30 | 1.10 | 25.00 | 0.10 | SLC12A3 | compound het | fs_ins | E78fs | - | - | 0.00E+00 | . | Dmis | R642H | D | 35 | 0 | Pathogenic |
| GIT466-1 | - | - | - | - | - | SLC12A3 | compound het | Dmis | L850P | D | 26.3 | 0.0001 | Pathogenic | Dmis | A313V | T | 23 | 2.03E-05 | Pathogenic |
| GIT478-1 | 107/64 | 2.60 | 0.80 | 28.00 | - | SLC12A3 | compound het | Dmis | G463R | D | 33 | 2.85E-05 | Pathogenic | Dmis | R958W | D | 28.4 | 0 | . |
| GIT483-1 | - | - | - | - | - | SLC12A3 | compound het | Dmis | G439S | D | 32 | 0.0002 | Pathogenic | Dmis | T649M | D | 32 | 3.67E-05 | Pathogenic |
| GIT486-1 | 110/70 | 3.10 | 1.30 | 24.00 | 0.07 | SLC12A3 | compound het | Dmis | L859P | D | 26.3 | 0.0001 | Pathogenic | Dmis | C994Y | D | 25.8 | 0.0002 | . |
| GIT494-1 | - | 2.40 | - | 29.60 | 0.26 | SLC12A3 | compound het | splicing | c.1180+1G>T | - | 35 | 2.80E-05 | Pathogenic | Dmis | P643L | D | 34 | 0.0002 | Pathogenic |
| GIT496-1 | 90/45 | 2.60 | 1.63 | 28.00 | - | SLC12A3 | compound het | splicing | c.1180+1G>T | - | 35 | 2.80E-05 | Pathogenic | Dmis | V647M | D | 32 | . | . |
| GIT505-1 | - | - | - | - | - | SLC12A3 | compound het | Dmis | G439S | D | 32 | 0.0002 | Pathogenic | Dmis | G729V | D | 27.6 | 0.0001 | Pathogenic |
| GIT513-1 | 96/56 | 1.60 | 1.20 | 28.00 | 0.00 | SLC12A3 | compound het | Dmis | R135C | T | 34 | 5.81E-05 | Pathogenic | stopgain | W161X | - | 38 | 1.23E-05 | Pathogenic |
| GIT524-1 | - | 2.60 | 1.40 | 25.00 | 0.04 | SLC12A3 | compound het | Dmis | G439S | D | 32 | 0.0002 | Pathogenic | Dmis | R955Q | T | 27.7 | 0.0002 | Pathogenic |
| GIT531-1 | 92/60 | 2.90 | 1.63 | 26.00 | 0.05 | SLC12A3 | compound het | Dmis | C994Y | D | 25.7 | 1.90E-04 | . | CNV | ex4-5 del | - | - | 0 | . |
| GIT537-1 | 134/100 | 3.80 | 1.20 | - | - | SLC12A3 | compound het | Dmis | A313V | T | 23 | 2.03E-05 | Pathogenic | Dmis | V142L | D | 29.6 | 4.34E-06 | . |
| GIT538-1 | - | - | - | - | - | SLC12A3 | compound het | Dmis | G741R | D | 27.5 | 4.00E-04 | Pathogenic | Dmis | S555L | D | 34 | 6.50E-05 | Pathogenic |

### Table S5 (cont.)

| PID | BP | $K^+$ | $Mg^{2+}$ | $HCO_3^-$ | $UCa^{2+}$:Cr | gene | genotype | mutation #1 type | mutation #1 | Meta SVM | CADD20 | gnomAD WES | ClinVar | mutation #2 type | mutation #2 | Meta SVM | CADD20 | gnomAD WES | ClinVar |
|---|---|---|---|---|---|---|---|---|---|---|---|---|---|---|---|---|---|---|---|
| GIT544-1 | - | - | - | - | - | SLC12A3 | compound het | Dmis | L859P | D | 26.3 | 0.0001 | Pathogenic | fs_ins | V398fs | - | - | 1.24E-05 | . |
| GIT545-1 | - | - | - | - | - | SLC12A3 | compound het | Dmis | G989R | T | 25.9 | 5.29E-05 | Pathogenic | Dmis | L272P | D | 28.4 | 1.63E-05 | Pathogenic |
| GIT565-1 | 104/54 | 3.20 | 1.10 | 29.00 | 0.05 | SLC12A3 | compound het | Dmis | C994Y | D | 25.7 | 1.90E-04 | . | CNV | ex24-25 del | - | - | 0 | . |
| GIT571-1 | - | - | - | - | - | SLC12A3 | compound het | splicing | c.2856+1G>T | - | 24.3 | 0.0002 | . | Dmis | R655C | D | 35 | 2.85E-05 | Pathogenic |
| GIT574-1 | 116/76 | 3.19 | 1.60 | 29.00 | 0.50 | SLC12A3 | compound het | Dmis | G989R | T | 25.9 | 5.29E-05 | Pathogenic | Dmis | Q1021R | T | 27 | 4.06E-06 | . |
| GIT589-1 | 130/80 | 2.64 | 1.97 | 28.00 | 0.05 | SLC12A3 | compound het | Dmis | R861C | D | 28 | 9.76E-05 | Pathogenic | Dmis | R955W | T | 27.7 | 4.06E-05 | Pathogenic |
| GIT610-1 | - | 2.90 | 0.93 | 32.00 | - | SLC12A3 | compound het | Dmis | R642G | D | 33 | 2.64E-05 | Pathogenic | fs_del | S126fs | - | - | 0 | . |
| GIT623-1 | - | - | - | - | - | SLC12A3 | compound het | fs_ins | E78fs | - | - | 0.00E+00 | . | Dmis | R135C | T | 34 | 5.81E-05 | Pathogenic |
| GIT627-1 | 134/80 | 2.30 | 1.10 | 34.00 | 0.88 | SLC12A3 | compound het | Dmis | G741R | D | 27.5 | 4.00E-04 | Pathogenic | Dmis | A298D | T | 28.8 | 0 | . |
| GIT633-1 | 120/70 | 2.40 | - | - | - | SLC12A3 | compound het | Dmis | G729V | D | 27.6 | 0.0001 | Pathogenic | stopgain | R1009X | - | 44 | 8.12E-06 | Pathogenic |
| GIT634-1 | 130/78 | 1.80 | - | - | - | SLC12A3 | compound het | Dmis | G439S | D | 32 | 0.0002 | Pathogenic | splicing | c.2856+1G>T | - | 24.3 | 0.0002 | . |
| GIT640-1 | 102/58 | 4.00 | 1.75 | 28.00 | - | SLC12A3 | compound het | splicing | c.965-1G>A | - | 24.5 | 0 | . | Dmis | I324N | T | 29.6 | . | . |
| GIT642-1 | 130/64 | 2.30 | 1.30 | 27.00 | 0.08 | SLC12A3 | compound het | Dmis | G439S | D | 32 | 0.0002 | Pathogenic | Dmis | G731R | D | 32 | 2.03E-05 | . |
| GIT653-1 | 105/70 | 3.00 | - | - | - | SLC12A3 | compound het | Dmis | G741R | D | 27.5 | 4.00E-04 | Pathogenic | Dmis | R852C | D | 28 | 9.76E-05 | Pathogenic |
| GIT689-1 | 111/56 | 2.60 | 1.80 | 24.00 | - | SLC12A3 | compound het | Dmis | T649M | D | 32 | 3.70E-05 | Pathogenic | fs_ins | T182fs | D | 32 | 3.67E-05 | . |
| GIT695-1 | 88/55 | 3.10 | 1.70 | 30.00 | - | SLC12A3 | compound het | Dmis | G741R | D | 27.5 | 4.00E-04 | Pathogenic | splicing | c.1180+1G>T | - | 26.4 | 2.87E-05 | . |
| GIT702-1 | 106/70 | 3.50 | 2.30 | 30.00 | - | SLC12A3 | compound het | Dmis | T382M | D | 34 | 5.32E-05 | Pathogenic | Dmis | W568R | D | 25 | 8.13E-06 | . |
| GIT705-1 | 114/70 | 2.90 | 1.60 | 36.00 | - | SLC12A3 | compound het | Dmis | G989R | T | 25.9 | 5.29E-05 | Pathogenic | Dmis | L177P | D | 25.1 | . | . |
| GIT706-1 | 110/70 | 2.40 | - | 29.00 | 0.03 | SLC12A3 | compound het | stopgain | R1018X | - | 44 | 8.12E-06 | Pathogenic | Dmis | L858H | D | 27.4 | 6.51E-05 | Pathogenic |
| GIT727-1 | 121/69 | 2.90 | 1.30 | 28.00 | 0.05 | SLC12A3 | compound het | Dmis | G741R | D | 27.5 | 4.00E-04 | Pathogenic | Dmis | S178L | D | 28.2 | 1.63E-05 | Pathogenic |
| GIT734-1 | 104/53 | 2.60 | 1.90 | 35.00 | - | SLC12A3 | compound het | Dmis | G741R | D | 27.5 | 4.00E-04 | Pathogenic | Dmis | R887Q | D | 34 | 4.47E-05 | Pathogenic |
| GIT614-1 | - | 2.90 | 1.30 | 30.00 | 0.50 | SLC12A3 | compound het | Dmis | T60M | D | 27.1 | 6.10E-05 | Pathogenic | 6-bp duplication | P643Q644ins | - | - | 4E-06 | - |
| GIT460-1 | 120/78 | 3.00 | 1.50 | 31.00 | 1.00 | SLC12A3 | compound het | fs_ins | A166fs | . | . | 0.00E+00 | . | CNV | del_exons_4-6 | - | - | 0 | - |
| GIT563-1 | 107/90 | 2.90 | 1.20 | 28.00 | 0.70 | SLC12A3 | compound het | splicing | c.602-2A>G | . | 22.8 | 1.00E-04 | . | CNV | del_exons_2-5 | - | - | 0 | - |
| GIT691-1 | 90/54 | 1.90 | 1.81 | 29.00 | - | SLC12A3 | compound het | Dmis | R852C | D | 28 | 9.80E-05 | Pathogenic | CNV | del_exons_1-7 | - | - | 0 | - |
| GIT723-1 | - | - | - | - | - | SLC12A3 | compound het | splicing | c.2856+1G>T | . | 24.3 | 2.00E-04 | . | CNV | ex2-3 del | - | - | 0 | - |
| GIT741-1 | 90/39 | 2.20 | 1.60 | 28.00 | 0.03 | SLC12A3 | compound het | Dmis | A313V | T | 23 | 2.00E-05 | Pathogenic | CNV | del_exons_24. 25 | - | - | 0 | - |
| GIT741-2 | 100/51 | 2.60 | 1.70 | 28.00 | - | SLC12A3 | compound het | Dmis | A313V | T | 23 | 2.00E-05 | Pathogenic | CNV | del_exons_24. 25 | - | - | 0 | - |
| GIT398-1 | 120/75 | 2.80 | 1.22 | 35.00 | 0.09 | SLC12A3 | compound het | splicing | c.2856+1G>T | . | 24.3 | 2.00E-04 | . | splice loss | c.2948+5 AG>AA | - | 24.3 | 0 | - |
| GIT418-1 | 120/80 | 3.20 | 1.56 | 31.10 | - | SLC12A3 | compound het | stopgain | R1009X | . | 44 | 8.12E-06 | Pathogenic | splice loss | c.2952-9 AT>AA | - | 22.5 | 0 | - |
| GIT699-1 | 116/63 | 2.10 | 1.80 | 36.00 | - | SLC12A3 | compound het | Dmis | G989R | T | 25.9 | 5.30E-05 | Pathogenic | splice loss | c.2037+4A>G | - | 23.7 | 4E-06 | - |

### Table S5 (cont.)

| PID | BP | K+ | Mg2+ | HCO3- | UCa2+/Cr | age at presentation | gene | genotype | mutation #1 type | mutation #1 | Meta SVM | CADD20 | gnomAD WES | ClinVar | mutation #2 type | mutation #2 | gnomAD WGS |
|---|---|---|---|---|---|---|---|---|---|---|---|---|---|---|---|---|---|
| GIT526-1 | 90/50 | 2.40 | 1.30 | 30.00 | 0.16 | 6 | SLC12A3 | monoallelic | Dmis | L859P | D | 26.3 | 0.0001 | Pathogenic | CNV | ex2-3 del | 0 |
| GIT336-1 | 140/90 | 2.60 | 1.20 | 32.00 | 0.02 | 10 | SLC12A3 | monoallelic | stopgain | R896X | . | 44 | 4.00E-04 | Pathogenic | regulatory mutation | 16-56900664 G>A | 0 |
| GIT250-1 | 88/54 | 2.20 | 1.50 | 28.00 | 0.08 | 9 | SLC12A3 | monoallelic | Dmis | T649M | D | 32 | 3.67E-05 | Pathogenic | splice gain | c.1670-191 GC>GT | 3.20E-05 |
| GIT255-1 | 114/74 | 2.60 | 0.87 | 31.00 | 0.07 | 41 | SLC12A3 | monoallelic | Dmis | L858H | D | 27.4 | 6.51E-05 | Pathogenic | splice gain | c.1670-191 GC>GT | 3.20E-05 |
| GIT125-1 | 120/80 | 2.40 | 1.71 | 27.00 | 0.03 | 28 | SLC12A3 | monoallelic | fs_ins | F765fs | - | - | 0 | . | WT | WT | - |
| GIT129-1 | 110/70 | 2.60 | 1.30 | 30.50 | 0.47 | 11 | SLC12A3 | monoallelic | Dmis | R861C | D | 28 | 8.50E-05 | Pathogenic | WT | WT | - |
| GIT420-1 | 132/80 | 3.10 | 1.50 | 25.00 | 0.09 | 47 | SLC12A3 | monoallelic | splicing | c.2856+1G>T | . | 24.3 | 2.00E-04 | . | WT | WT | - |

**Table S6. List of probands with monoallelic coding damaging missense or loss of function mutations in SLC12A3 and noncoding mutations identified by WGS.** Clinical information, followed by information on the biallelic mutations, is shown.

| PID | BP | $K^{+}$ | $Mg^{2+}$ | $HCO3^{-}$ | $UCa^{2+}$:Cr | gene #1 | genotype | mutation #1 type | mutation #1 | MetaSVM | CADD20 | gnomAD WES | ClinVar | mutation #2 type | mutation #2 | Meta SVM | CADD20 | gnomAD WES | ClinVar |
|---|---|---|---|---|---|---|---|---|---|---|---|---|---|---|---|---|---|---|---|
| GIT171-1 | - | - | - | - | - | CLCNKB | homozygote | CNV | 12.9kb_del | - | - | 0 | | - | - | - | - | - | - |
| GIT311-1 | 136/68 | 3.60 | 1.70 | 32.00 | 0.14 | CLCNKB | homozygote | stopgain | R595X | - | 36 | 9.70E-05 | Pathogenic | - | - | - | - | - | - |
| GIT331-1 | - | - | - | - | - | CLCNKB | homozygote | fs_del | S445fs | - | - | 0 | . | - | - | - | - | - | - |
| GIT490-1 | 100/20 | 2.80 | 1.82 | 30.00 | 0.18 | CLCNKB | homozygote | Dmis | A242E | D | 32 | 4.00E-06 | . | - | - | - | - | - | - |
| GIT493-1 | 80/20 | 2.50 | 1.95 | 32.00 | 2.52 | CLCNKB | homozygote | stopgain | R595X | - | 36 | 2.80E-05 | Pathogenic | - | - | - | - | - | - |
| GIT495-1 | 90/50 | 2.20 | 1.95 | 33.00 | 2.00 | CLCNKB | homozygote | CNV chimera | 12.4kb_del | - | - | 0 | | - | - | - | - | - | - |
| GIT536-1 | - | 3.17 | 2.43 | 34.50 | 0.39 | CLCNKB | homozygote | CNV | 21.4kb_del | - | - | 0 | | - | - | - | - | - | - |
| GIT539-1 | - | - | - | - | - | CLCNKB | homozygote | CNV | 12.9kb_del | - | - | 0 | - | - | - | - | - | - | - |
| GIT540-1 | - | - | - | - | - | CLCNKB | homozygote | stopgain | R663X | - | 37 | 0 | . | - | - | - | - | - | - |
| GIT546-1 | - | - | - | - | - | CLCNKB | homozygote | CNV | 12.9kb_del | - | - | 0 | - | - | - | - | - | - | - |
| GIT551-1 | - | - | - | - | - | CLCNKB | homozygote | Dmis | G437R | D | 32 | 2.90E-05 | Pathogenic | - | - | - | - | - | - |
| GIT553-1 | - | 2.80 | 2.30 | 32.00 | - | CLCNKB | homozygote | CNV | 12.9kb_del | - | - | 0 | | - | - | - | - | - | - |
| GIT554-1 | 130/76 | 3.00 | 1.90 | 32.20 | 0.80 | CLCNKB | homozygote | Dmis | A204T | D | 26.4 | 7.40E-05 | Pathogenic | - | - | - | - | - | - |
| GIT566-1 | 91/60 | 3.20 | 1.40 | - | - | CLCNKB | homozygote | Dmis | V170M | D | 28.7 | 1.00E-04 | . | - | - | - | - | - | - |
| GIT573-1 | 110/70 | 2.80 | 1.50 | 32.00 | 0.03 | CLCNKB | homozygote | nfs_del | del397-400 | - | - | 4.00E-06 | . | - | - | - | - | - | - |
| GIT578-1 | 130/80 | 2.80 | 2.00 | 38.00 | 0.30 | CLCNKB | homozygote | Dmis | V170M | D | 28.7 | 1.00E-04 | . | - | - | - | - | - | - |
| GIT579-1 | 86/46 | 2.40 | - | 49.00 | 1.50 | CLCNKB | homozygote | Dmis | R438C | D | 35 | 2.44E-05 | . | - | - | - | - | - | - |
| GIT582-1 | - | - | - | - | - | CLCNKB | homozygote | Dmis | A204T | D | 26.4 | 7.40E-05 | Pathogenic | - | - | - | - | - | - |
| GIT595-1 | 98/50 | 2.07 | 2.00 | 28.00 | - | CLCNKB | homozygote | CNV | 12.9kb_del | - | - | 0 | | - | - | - | - | - | - |
| GIT602-1 | 103/60 | 2.10 | 2.20 | 30.00 | 0.72 | CLCNKB | homozygote | CNV | 12.9kb_del | - | - | 0 | | - | - | - | - | - | - |
| GIT605-1 | 90/60 | 2.50 | 1.70 | 59.90 | 0.54 | CLCNKB | homozygote | Dmis | P124L | D | 27.4 | 1.20E-05 | Pathogenic | - | - | - | - | - | - |
| GIT606-1 | 100/65 | 2.30 | - | - | - | CLCNKB | homozygote | CNV | 12.9kb_del | - | - | 0 | | - | - | - | - | - | - |
| GIT612-1 | - | - | - | - | - | CLCNKB | homozygote | splicing | c.274+2T>A | - | 24 | 0 | . | - | - | - | - | - | - |
| GIT618-1 | - | - | - | - | - | CLCNKB | homozygote | CNV | 12.9kb_del | - | - | 0 | | - | - | - | - | - | - |
| GIT631-1 | - | 2.60 | - | 38.00 | 1.00 | CLCNKB | homozygote | fs_del | T632fs | - | - | 7.70E-05 | Pathogenic | - | - | - | - | - | - |
| GIT677-1 | 74/52 | 2.10 | - | 30.00 | - | CLCNKB | homozygote | CNV | 1.26 Mb del | - | - | 0 | | - | - | - | - | - | - |
| GIT701-1 | - | 2.20 | 1.80 | 32.00 | - | CLCNKB | homozygote | Dmis | W96R | D | 24.8 | 0 | . | - | - | - | - | - | - |
| GIT712-1 | 90/60 | 3.20 | 1.20 | - | - | CLCNKB | homozygote | CNV | 12.9kb_del | - | - | 0 | | - | - | - | - | - | - |
| GIT729-1 | - | 2.25 | - | - | - | CLCNKB | homozygote | Dmis | A469P | D | 27.4 | 0 | . | - | - | - | - | - | - |
| GIT542-1 | - | - | - | - | - | CLCNKB | compound het | Dmis | G296D | D | 13.26 | 0 | . | splicing | c.1419+1G>A | - | 23.9 | 0 | . |
| GIT548-1 | - | - | - | - | - | CLCNKB | compound het | fs_del | L564fs | - | - | 6.40E-05 | - | splicing | c.461+1G>A | - | 33 | 8.00E-06 | . |
| GIT569-1 | 112/65 | 3.30 | 2.20 | 33.00 | 1.00 | CLCNKB | compound het | Dmis | V170M | D | 28.7 | 0.0001 | . | Dmis | R438C | D | 35 | 2.44E-05 | . |
| GIT598-1 | - | - | - | - | - | CLCNKB | compound het | Dmis | R438C | D | 35 | 2.44E-05 | . | Dmis | R438H | D | 34 | 2.03E-05 | . |
| GIT619-1 | 118/58 | 2.30 | 2.60 | 32.10 | - | CLCNKB | compound het | splice donor loss | c.100+2GT>G | - | - | 0 | . | stopgain | W610X | - | 39 | 0.0002 | . |
| GIT636-1 | 120/80 | 2.40 | - | - | - | CLCNKB | compound het | Dmis | A204T | D | 26.4 | 7.40E-05 | Pathogenic | stopgain | R595X | - | 36 | 0.00009587 | Pathogenic |
| GIT648-1 | 134/84 | 2.80 | 1.90 | 26.00 | - | CLCNKB | compound het | stopgain | R304X | - | 41 | 4.00E-05 | Pathogenic | CNV | ex12-13 del | - | - | 0 | - |

**Table S7. List of probands with biallelic mutations in CLCNKB, BSND, CLDN10, KCNJ1, KCNJ10, MAGED2 SLC12A1, and SLC26A3 in the salt-wasting cohort.** Clinical information, followed by information on the biallelic mutations, is shown.

| PID | BP | $K^+$ | $Mg^{2+}$ | $HCO_3^-$ | $UCa^{2+}$:Cr | age at diagnosis | gene #1 | genotype | mutation #1 type | mutation #1 | Meta SVM | CADD20 | gnomAD WES | ClinVar | mutation #2 type | mutation #2 | Meta SVM | CADD20 | gnomAD WES | ClinVar |
|---|---|---|---|---|---|---|---|---|---|---|---|---|---|---|---|---|---|---|---|---|
| GIT128-1 | 100/64 | 2.80 | 2.20 | 29.00 | 0.00 | 44 | CLDN10 | homozygote | Dmis | C128R | D | 25.7 | 0 | . | - | - | - | - | - | - |
| GIT182-1 | 110/70 | 1.90 | 1.87 | 24.00 | 1.10 | 0 | SLC12A1 | homozygote | Dmis | Y528D | D | 28 | 0 | . | - | - | - | - | - | - |
| GIT191-1 | 96/60 | 3.50 | 1.87 | 28.00 | - | 0 | KCNJ1 | homozygote | CNV | ex1-2 | - | - | 0.00E+00 | | - | - | - | - | - | - |
| GIT257-1 | 110/59 | 2.80 | 2.31 | 32.00 | 1.20 | 0.5 | KCNJ1 | homozygote | CNV | ex1-2 | - | - | 0 | | - | - | - | - | - | - |
| GIT287-1 | 77/38 | 2.70 | 1.30 | 29.00 | 1.92 | 0 | SLC12A1 | homozygote | CNV | ex10del | - | - | 0.00E+00 | | - | - | - | - | - | - |
| GIT326-1 | 110/75 | 2.80 | 2.00 | 38.00 | 0.04 | 13 | CLDN10 | homozygote | Dmis | P147L | D | 29.5 | 0.00E+00 | . | - | - | - | - | - | - |
| GIT396-1 | - | - | - | - | - | - | SLC26A3 | homozygote | Dmis | D652N | D | 34 | 4.00E-06 | . | - | - | - | - | - | - |
| GIT485-1 | 78/- | 4.70 | 0.86 | 22.00 | 2.20 | 0 | KCNJ1 | homozygote | stopgain | W80X | - | 36 | 0 | . | - | - | - | - | - | - |
| GIT492-1 | - | - | - | - | - | - | KCNJ1 | homozygote | Dmis | H251R | D | 24.5 | 0 | . | - | - | - | - | - | - |
| GIT532-1 | - | 3.10 | 1.70 | 22.00 | 0.40 | 2 | KCNJ1 | homozygote | Dmis | A195V | D | 23.9 | 1.60E-05 | . | - | - | - | - | - | - |
| GIT594-1 | - | - | - | - | - | 2.5 | KCNJ10 | homozygote | Dmis | R65P | D | 28.5 | 0.00E+00 | Pathogenic | - | - | - | - | - | - |
| GIT609-1 | - | 6.50 | 3.00 | 24.00 | - | 0 | MAGED2 | homozygote | Dmis | R446P | T | 32 | 0.00E+00 | . | - | - | - | - | - | - |
| GIT644-1 | - | - | - | - | - | 0.2 | BSND | homozygote | fs_del | G150fs | - | - | 4.10E-05 | . | - | - | - | - | - | - |
| GIT647-1 | - | - | - | - | - | - | KCNJ1 | homozygote | Dmis | T51I | D | 25 | 8.10E-06 | . | - | - | - | - | - | - |
| GIT649-1 | - | - | - | - | - | 2 | KCNJ1 | homozygote | Dmis | L340R | D | 23.3 | 1.00E-04 | . | - | - | - | - | - | - |
| GIT651-1 | - | - | - | - | - | - | SLC12A1 | homozygote | fs_del | E993fs | - | - | 0.00E+00 | Pathogenic | - | - | - | - | - | - |
| GIT654-1 | 108/84 | 4.20 | 1.90 | 23.90 | - | 0 | MAGED2 | homozygote | Dmis | R446C | T | 34 | 0 | Pathogenic | - | - | - | - | - | - |
| GIT656-1 | 90/60 | 3.30 | - | - | - | 3 | SLC12A1 | homozygote | nfs_del | del525-526 | - | - | 0 | . | - | - | - | - | - | - |
| GIT671-1 | - | - | - | - | - | - | SLC26A3 | homozygote | Dmis | V560D | D | 34 | 0 | . | - | - | - | - | - | - |
| GIT684-1 | - | - | - | - | - | 0 | SLC12A1 | homozygote | Dmis | G397D | D | 30 | 0 | . | - | - | - | - | - | - |
| GIT783-1 | - | 3.60 | 0.64 | 29.00 | - | 35 | MT-TI | homoplasmic | - | - | - | - | 0 | | - | - | - | - | - | - |
| GIT270-1 | 106/67 | 4.40 | 2.00 | 20.00 | 1.40 | 0.75 | SLC12A1 | compound het | Dmis | R199C | D | 33 | 4.00E-06 | . | stopgain | Q656X | . | 46 | 8.30E-06 | . |
| GIT296-1 | 91/45 | 3.30 | 1.10 | 24.70 | 2.80 | 0 | BSND | compound het | Dmis | M1I | D | 27.6 | 4.06E-05 | Pathogenic | Dmis | G10S | D | 32 | 4.06E-06 | Pathogenic |
| GIT332-1 | 99/66 | 2.30 | 2.82 | 26.00 | 4.20 | 4 | BSND | compound het | Dmis | G47R | T | 28.3 | 9.77E-05 | Pathogenic | LOF | P134fs | - | - | 0 | Pathogenic |
| GIT473-1 | 94/57 | 4.90 | 1.80 | 22.00 | 2.30 | 0 | SLC12A1 | compound het | stopgain | Y639X | - | 35 | 0 | Pathogenic | splicing | c.2873+1G >A | - | 24.7 | 1.97E-05 | Pathogenic |
| GIT528-1 | 90/51 | 5.20 | 2.60 | 26.00 | - | 0 | SLC12A1 | compound het | Dmis | G397A | D | 27.2 | 4.07E-06 | . | splicing | c.3164+1G >A | - | 27 | 5.02E-06 | Pathogenic |
| GIT591-1 | 109/60 | 3.30 | 1.78 | 29.00 | - | 0 | SLC12A1 | compound het | Dmis | R199H | D | 35 | 0 | . | Dmis | A515S | D | 32 | 0 | . |
| GIT596-1 | 119/58 | 2.50 | 1.70 | 26.00 | - | 35 | SLC12A1 | compound het | Dmis | R439Q | D | 35 | 2.03E-05 | Pathogenic | Dmis | F611Y | D | 24.6 | 0 | . |
| GIT703-1 | 64/36 | 4.60 | 1.50 | 26.00 | - | 0 | SLC12A1 | compound het | LOF | S120Rfs*15 | - | - | 0 | - | splicing | c.629-6A >G | - | 16.2 | 0 | - |
| GIT615-1 | - | 4.90 | 1.21 | 25.00 | - | 3 | SLC12A1 | compound het | Dmis | P544S | D | 29.8 | 0 | . | splicing | c.2402+1G >A | - | 24.8 | 8.18E-06 | . |
| GIT725-1 | - | - | - | - | - | 4 | SLC12A1 | compound het | fs_ins | R116fs | - | - | 0 | . | Dmis | Y246C | D | 28.8 | 2.90E-05 | . |
| GIT739-1 | 60/40 | 2.80 | 1.30 | 30.00 | - | 0 | SLC12A1 | compound het | splicing | c.3164+1G>A | - | 27 | 5.02E-06 | . | splicing | c.1560+1G >A | - | 26.4 | 2.45E-05 | . |

### Table S7 (cont.)
